# Mucosal and Systemic Immune Correlates of Viral Control following SARS-CoV-2 Infection Challenge in Seronegative Adults

**DOI:** 10.1101/2023.07.21.23292994

**Authors:** Helen R. Wagstaffe, Ryan S. Thwaites, Arnold Reynaldi, Jasmin K. Sidhu, Richard McKendry, Stephanie Ascough, Loukas Papargyris, Ashley M. Collins, Jiayun Xu, Nana-Marie Lemm, Matthew K. Siggins, Benny M. Chain, Ben Killingley, Mariya Kalinova, Alex Mann, Andrew Catchpole, Miles P. Davenport, Peter J.M. Openshaw, Christopher Chiu

**Affiliations:** Department of Infectious Disease, Imperial College London, London, United Kingdom; National Heart and Lung Institute, Imperial College London, London, United Kingdom; Kirby Institute, University of New South Wales, Kensington, NSW, Australia; UCL Division of Infection and Immunity, University College London, London, UK; Department of Infectious Diseases, University College London Hospital, London, UK; hVIVO Services Ltd., London, UK

## Abstract

Human infection challenge permits characterisation of the associated immune response in unparalleled depth, enabling evaluation of early pre-symptomatic immune changes and the dynamic immune factors important for viral clearance. Here, 34 healthy young adult volunteers, seronegative to SARS-CoV-2, were inoculated with a D614G-containing pre-Alpha SARS-CoV-2 strain. Nasal and systemic soluble mediator and antibody responses, and peripheral blood T cell and B cell responses were measured by MesoScale Discovery and flow cytometry just before and up to 1 year after intra-nasal inoculation. In the 18 (53%) participants who became infected, both nasal and systemic mediator responses were dominated by interferons (IFN) but with divergent kinetics. T cell activation and proliferation in blood peaked at day 10 in CD4^+^ T cells and day 14 in CD8^+^ T cells, returning to baseline by day 28. Following infection, antigen-specific T cells were largely CD38^+^Ki67^+^ and displayed central and effector memory phenotypes. T cells contracted after viral clearance with expanded antigen-specific memory T cell populations persisting past day 28. Both mucosal and systemic antibodies became detectable around day 10 but nasal antibodies plateaued after day 14 while circulating antibodies continued to rise. Using piecewise linear regression modelling, viral load related closely to the induction of type I IFN responses, moreover, CD8^+^ T cell responses and early IgA responses were strongly associated with viral clearance. Detailed analysis of innate and adaptive immune responses to primary SARS-CoV-2 infection following human challenge thus revealed the relationship between immune kinetics and viral load as factors associated with resolution of infection.

## Introduction

SARS-CoV-2 has caused more than 6 million documented deaths from COVID-19 since its emergence in late 2019^1^, with estimates of excess mortality resulting from the pandemic as high as 18 million. Vaccines and therapeutics that inhibit the inflammatory response have effectively reduced severe outcomes^2, 3^, but new infections and transmission remain high even among vaccinated people due to waning immunity and limited cross-strain protection^4^. Further understanding of the human immune response to SARS-CoV-2 and how dynamic innate and adaptive immune responses influence viral replication, especially in the critical early pre-symptomatic stage of infection, is required to accelerate the development of more effective interventions^5^.

Observational studies in humans are poorly suited to studying the early pre-symptomatic period of respiratory virus infection and rapidly induced immune responses, particularly in mild cases that represent successful immune containment and elimination of infection. Unavoidable gaps in case ascertainment and unmeasurable confounders such as inoculum dose, strain and exposure limit interpretation of such data. Furthermore, difficulties in sampling the respiratory mucosa have restricted our understanding of immune responses at the earliest anatomical sites of SARS-CoV-2 replication. Human infection challenge (HIC) models enable standardisation of inoculation and precise alignment of immunological data to the known time of exposure. Such studies have previously enabled the identification of immune mechanisms associated with protection against infection and/or disease by viral pathogens, including the abundance of mucosal antibodies and early mucosal responses to exposure, along with the role of resident memory T cells in limiting disease severity^6–8^.

Here, we delineate innate and adaptive immune responses to SARS-CoV-2 human challenge in the respiratory mucosa and blood throughout the time-course of infection. Conducted early during the COVID-19 pandemic, this study provided the unique opportunity to characterise the primary immune response to a novel respiratory pathogen in seronegative young, healthy adults who either developed self-limiting PCR detectable infection with mild symptoms or remained uninfected following exposure to the same inoculum. The comprehensiveness and granularity of measurements allowed viral and immune response parameters to be determined with unprecedented accuracy and thus identification of the relationships between early immune responses in the mucosa and blood, viral replication and later cellular and humoral adaptive immunity. Using piecewise linear regression models to explain the interactive dynamics of measured parameters, we show how viral loads relate to the induction of type I interferon and the importance of CD8^+^ T cell and early IgA responses in viral clearance.

## Results

### Nose and throat are semi-independent sites of SARS-CoV-2 replication with differential viral load dynamics

Thirty-four healthy adult volunteers aged 18–29 years were inoculated with a wild-type D614G-containing pre-Alpha SARS-CoV-2 challenge virus as previously described^9^. Following inoculation, 18 (53%) developed sustained infection defined as 2 consecutive quantifiable viral load (VL) detections by qPCR in nose and/or throat swabs. Soluble mediator and antibody levels in nasal mucosal fluid sampled by nasosorption and plasma were measured daily and T and B cell responses in peripheral blood were measured at intervals before and after inoculation (Fig. 1a). The first 6 infected participants received pre-emptive anti-viral treatment with Remdesivir immediately after the first 2 consecutive PCR detections; there was no difference in VL or symptom scores between those treated with Remdesivir and the later groups ^9^ and these groups were therefore analysed together.

**Figure 1:**
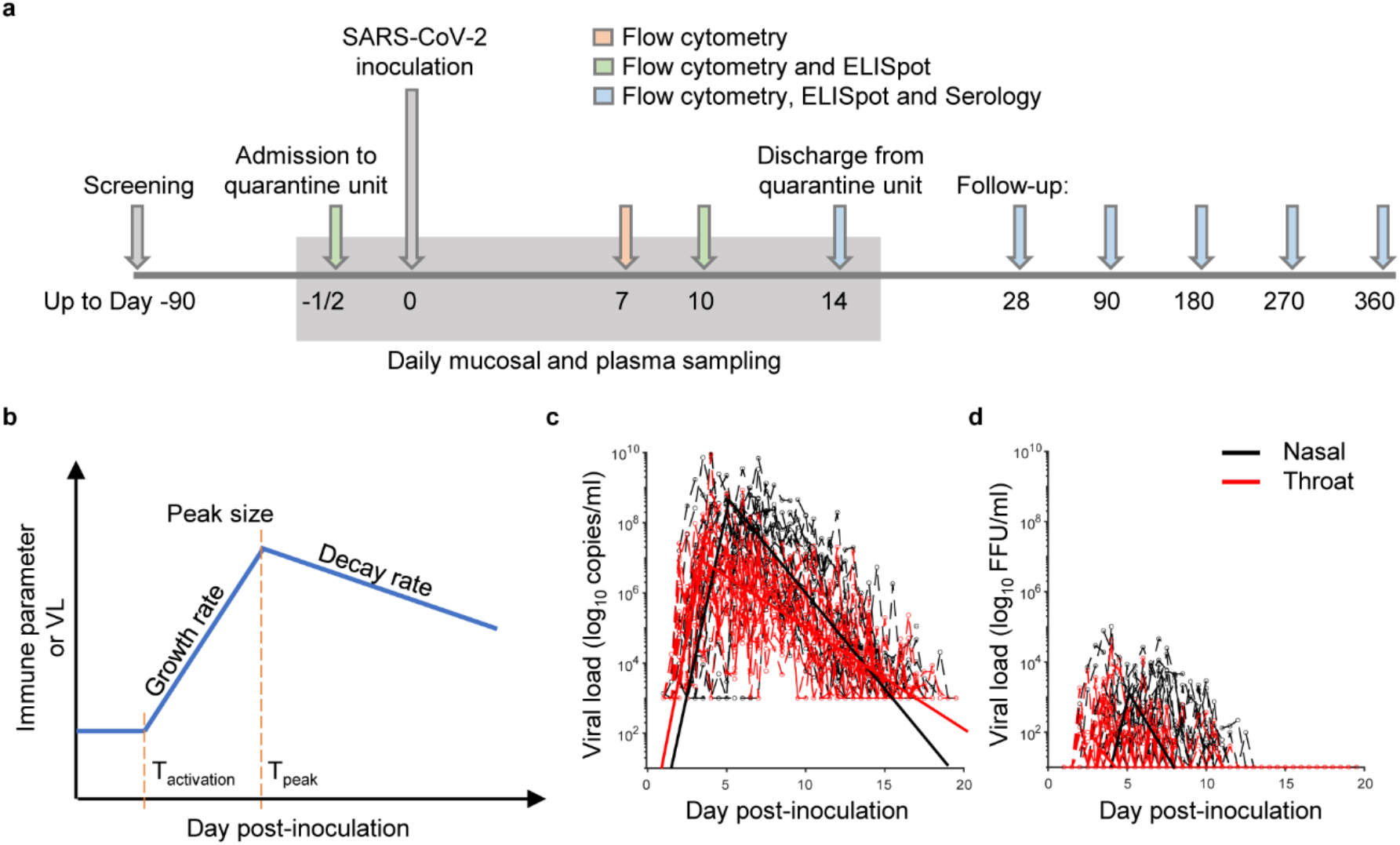
Study design and piecewise linear regression modelling of viral replication following SARS-CoV-2 human challenge. Thirty-four healthy adults aged 18-29 years with no evidence of previous SARS-CoV-2 vaccination or infection were inoculated with a GMP pre-Alpha challenge virus. (A) The schematic shows the study design, including sampling timepoints for mucosal and plasma soluble mediators and antibodies, flow cytometry and ELISpot assays. (B) Schematic showing piecewise linear regression model parameters and the model applied to (C) qPCR and (D) focus forming assay (FFA) viral load data from the nose and throat in participants with sustained infection (n=18).

Using piecewise linear regression models, VL and immune kinetics parameters including the exact time of onset and dynamic characteristics were estimated (Fig. 1b), defining kinetic parameters of the primary immune response to SARS-CoV-2 infection. The model assumed a constant initial level (or limit of detection for VL) until growth began (at T_activation_), constant exponential growth rate from T_activation_ to peak (T_peak_) and decay rate after the peak (Figure 1b). The model was applied to both qPCR and focus forming assay (FFA) VL measurements, although the low sensitivity of the FFA assay provided more limited data (Fig. 1c,d).

Our previous analysis of VL in those participants who became infected, showed earlier detection of viral replication in the throat compared with the nose^9^. This was confirmed by mathematical modelling (T_activation_ throat 1.78 days. vs nose 2.61 days; p=0.0054). The VL growth rate was similar between the throat and nose (throat 5.41 days^-1^ vs nose 4.86 days^-1^; ns) (Fig. 1c) but the T_peak_ was earlier in the throat than the nose (throat 3.4 days vs nose 5.1 days.; p<0.0001), with a lower peak VL (throat 6.96 log_10_ vs nose 8.69 log_10_; p<0.0001). However, the VL decay rate was lower in the throat (0.69 day^-1^ vs 1.29 day^-1^; p<0.0001) resulting in earlier viral clearance from the nose (Fig. 1c). Inter-individual variation in the throat VL data was significantly lower than in the nose, where there was marked variance in T_activation_ resulting in greater heterogeneity between individuals in nasal VL parameters. This was evidenced by the random effect on T_activation_, which was highly significant in the nose (p=3.04×10^-64^), but not significant in the throat (p=0.15, likelihood ratio test). We hypothesised that this heterogeneity reflected differences in immune responses between individuals, with differential impact on the nasal mucosa compared with the throat despite their anatomical proximity, implying that the two sites were semi-independently regulated.

### Nasal and systemic mediator responses are dominated by interferons

To characterise the nature and scale of the nasal mucosal and systemic soluble mediator responses following SARS-CoV-2 inoculation, a multiplexed panel of cytokines and chemokines was quantified in daily nasosorption and plasma samples. In infected participants, nasal IFN-α2a, IL-29 (IFN-λ) and IFN-y were significantly elevated compared with the uninfected group from day 4, followed by the IFN-induced chemokine CXCL10, CCL3, IL-15, and CCL4 at day 5 and CCL2 at day 6 (all p<0.05; Fig. 2a). These peaked between day 6 and 10 post-inoculation and then declined, with IL-29, IFN-y, CXCL10, CCL3, IL-15 and CCL4 still elevated at day 14 but returning to baseline by day 28 (Fig. 2a). There was no significant change in any other mediators measured in the nose (Supplementary Fig. 1a) and no change from baseline in the level of any of these proteins in the uninfected group. These mediators were additionally significantly different between infected and uninfected groups at different times over the study time course (Supplementary Video 1).

**Figure 2:**
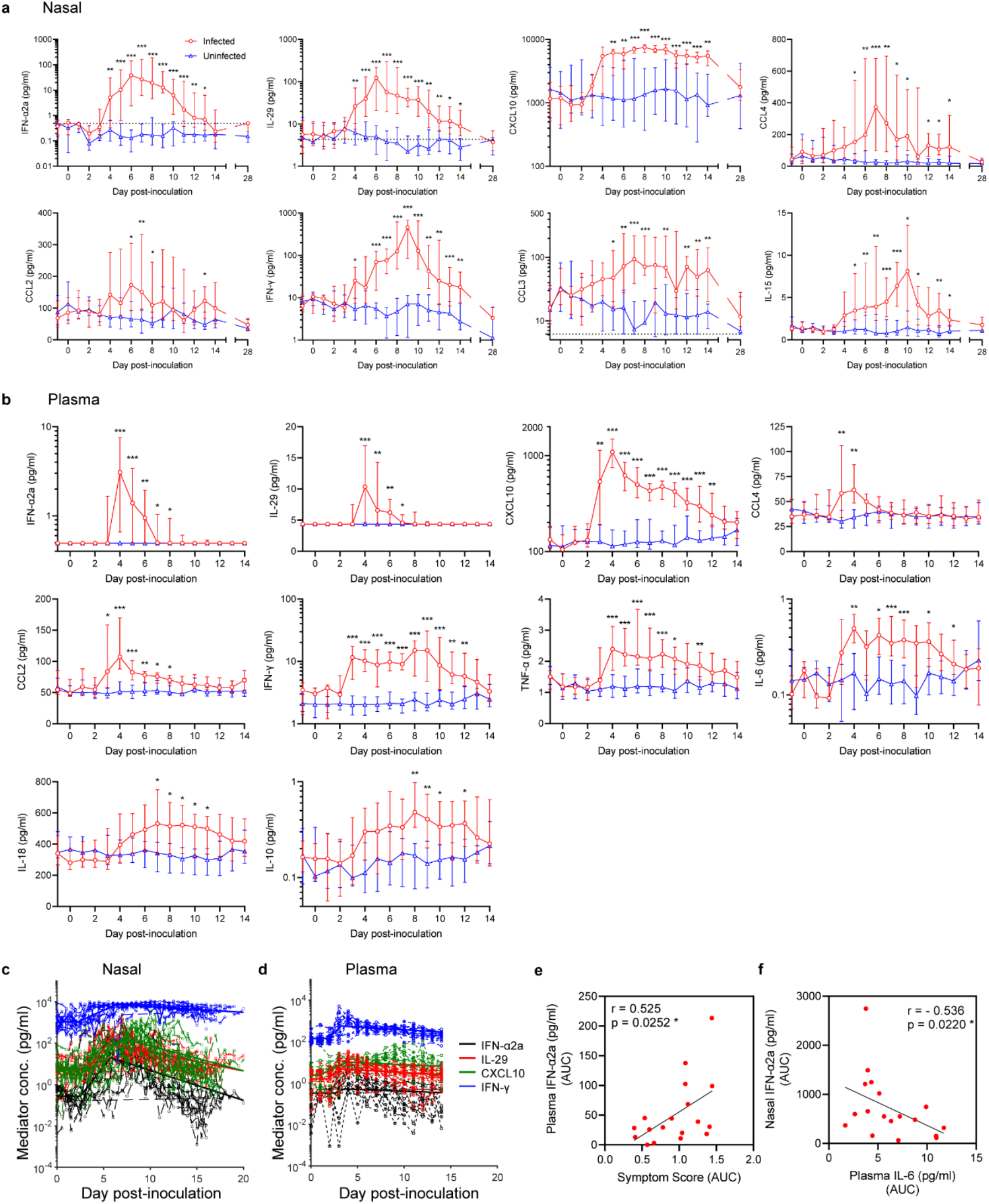
Interferon-dominated systemic inflammatory responses to SARS-CoV-2 precede those in the nasal mucosa. Soluble mediators in (A) nasal lining fluid and (B) plasma (B) were measured by MesoScale Discovery in infected (n=18) and uninfected individuals (n=16) after inoculation with SARS-CoV-2. Plots show medians and IQR. Significance between infected and uninfected groups was tested by multiple Mann-Whitney tests with Holm-Šídák’s correction for testing of multiple timepoints (adjusted *P* values *<0.05, **<0.01, ***<0.001). Participants vaccinated and infected in the community post-quarantine are excluded from day 28 data (see Supplementary Table 1 for details). IFN-α2a, IL-29, CXCL10 and IFN-y data were used for modelling the (C) nasal lining fluid and (D) plasma response. (E) Spearman correlation between plasma IFN-α2a and symptom score area under the curve (AUC) between days 0-14 and (F) plasma IFN-α2a and plasma IL-6 AUCs within the Infected group are shown.

Mediators detected in the plasma were similarly dominated by interferons; IFN-γ and the interferon-stimulated chemokine CXCL10 were significantly elevated over the uninfected group by day 3, IFN-α2a and IL-29 by day 4 (all p<0.05). CCL2, CCL4, TNF-α and IL-6 were also induced by day 3 and 4 (Fig. 2b). These remained elevated beyond day 6 with the exception of CCL4, while significant elevation of IL-18 and the inhibitory cytokine IL-10 were seen later from day 8. By day 12 the systemic inflammatory response had largely resolved with only IFN-γ remaining elevated (Fig. 2b). Apart from a modest elevation of plasma IL-15 in the infected group at day 6 (compared with the uninfected group; p=0.023; Supplementary Fig. 1b) there was no significant change in other measured mediators (Supplementary Fig. 1b). No plasma mediators were elevated in the uninfected group (Fig. 2b).

Mutations in IFN signalling are associated with severe COVID-19^10^, with improved clinical outcomes in some trials of IFN-β-1b treatment^11^. We therefore used piecewise linear regression to model the physiological IFN responses to investigate their interaction with clinical outcomes (Fig. 2c,d and Table 1). Analysing the nose and plasma compartments separately, IFN-α2a, IL-29, CXCL10 and IFN-γ responses became activated around the same time with T_activation_ between day 2 and 4 post-inoculation and no significant differences between the interferons. These responses also peaked at a similar time. These data therefore show that IFN-dominated responses at the site of virus entry and systemically occur rapidly post-infection in a largely in-compartment coordinated manner.

**Table 1:** Modelled nasal and systemic IFN-induced CXCL10 and IFN response parameters. Mean and 95% CI.

| | IFN-α2a | | | IL-29 | | | CXCL10 | | | IFN-γ | | |
| --- | --- | --- | --- | --- | --- | --- | --- | --- | --- | --- | --- | --- |
|  | Nasal | Plasma | Nasal vs plasma (p value) | Nasal | Plasma | Nasal vs plasma (p value) | Nasal | Plasma | Nasal vs plasma (p value) | Nasal | Plasma | Nasal vs plasma (p value) |
| T<sub>activation</sub> (day) | 2.47<br>(1.96 - 3.1) | 2.58<br>(0.52 - 12.87) | 0.95 (ns) | 3.25<br>(2.64 - 4) | 1.79<br>(0.93 - 3.43) | 0.008 | 2.25<br>(1.88 - 2.7) | 1.87<br>(1.15 - 3.06) | 0.24 (ns) | 3.56<br>(2.79 - 4.53) | 0.90<br>(0.23 - 3.61) | 0.02 |
| Growth rate (day<sup>-1</sup>) | 1.31<br>(0.99 - 1.7) | 0.46<br>(0.00276 - 798.8) | 0.78 (ns) | 1.24<br>(0.97 - 1.56) | 0.74<br>(0.34 - 1.66) | 0.07 (ns) | 0.48<br>(0.34 - 0.64) | 1.01<br>(0.41 - 2.44) | 0.0006 | 1.04<br>(0.85 - 1.24) | 0.48<br>(0.19 - 1.22) | 0.04 |
| T<sub>peak</sub> (day) | 5.75 (5.3 - 6.25) | 3.57<br>(0.97 - 13.16) | 0.44 (ns) | 5.47<br>(4.85 - 6.17) | 3.21<br>(2.28 - 4.53) | 2.7x10<sup>-6</sup> | 6.30<br>(5.51 - 7.19) | 3.23<br>(2.31 - 4.5) | 6.5x10<sup>-11</sup> | 6.96<br>(6.18 - 7.83) | 3.91<br>(2.11 - 7.22) | 0.02 |
| Decay rate (day<sup>-1</sup>) | 0.34<br>(0.25 - 0.41) | 0.046<br>(0.009 - 0.14) | 0.0001 | 0.21<br>(0.19 - 0.23) | 0.046<br>(0.039 - 0.069) | <2.2x10<sup>-16</sup> | 0.064<br>(0.053 - 0.08) | 0.062<br>(0.037 - 0.11) | 0.78 (ns) | 0.28<br>(0.23 - 0.34) | 0.046<br>(0.014 - 0.16) | 0.0005 |
| Peak size (log10) | 2.51<br>(2.32 - 2.7) | 0.57<br>(0.33 - 0.81) | 3.9x10<sup>-14</sup> | 2.66<br>(2.53 - 2.79) | 1.07 (0.9 - 1.23) | <2.2x10<sup>-16</sup> | 3.92 (3.8 - 4.04) | 3.15<br>(3.12 - 3.18) | 4.8x10<sup>-14</sup> | 2.91<br>(2.78 - 3.05) | 1.46<br>(1.27 - 1.64) | 4.09x10<sup>-14</sup> |
| AUC (log10) | 2.69<br>(2.59 - 2.78) | 1.26<br>(1.09 - 1.43) | 7.6x10<sup>-16</sup> | 2.92<br>(2.83 - 3.02) | 1.78<br>(1.72 - 1.89) | <2.2x10<sup>-16</sup> | 4.87<br>(4.81 - 4.93) | 3.79<br>(3.75 - 3.85) | <2.2x10<sup>-16</sup> | 3.33<br>(3.32 - 3.35) | 2.23 (1 - 3.09) | 7x10<sup>-4</sup> |

We previously showed that there was no correlation between the magnitude of VL and symptom severity^9^ and, similarly, no correlations were found between the model derived VL parameters and symptom scores in this analysis. Symptom scores did however weakly correlate with systemic mediator responses, where there was a significant positive correlation with plasma IFN-α2a AUC (Fig. 2e). This suggests that symptoms experienced during mild SARS-CoV-2 infection may be more closely associated with immunological responses than VL.

### Nasal and circulating inflammatory responses are distinct in scale, timing, and breadth

Comparing the kinetics of nasal and plasma responses (Fig. 2d and Table 1), the T_activation_ of IFN-α2a and IL-29 were similar between compartments, but the IFN-stimulated CXCL10 and IFN-γ responses were upregulated significantly earlier in plasma. In contrast, nasal IFN-α2a, CXCL10 and IFN-γ all peaked later and remained elevated for longer than their systemic counterparts (Fig. 2 and Table 1). Nasal responses were also higher in magnitude compared with the plasma by peak size and AUC (Table 1). Correlation matrix analysis demonstrated strong positive correlations between mediators such as the type I and II IFNs, IFN-α2a and IL-29, and myeloid cell chemoattractants, CCL3 and CCL4, within individual sites but few positive correlations between mediator responses by AUC between the nose and blood (Supplementary Fig. 2a). Conversely, a negative correlation was observed between nasal IFN-α2a and plasma IL-6 (Fig. 2f), suggesting that more robust local IFN responses might in part be associated with less marked peripheral inflammatory responses.

To determine the heterogeneity of nasal and plasma immune mediator responses between individuals, fold-change analyses (relative to day 0; baseline) at the median peak timepoint in the infected group for each of these mediators was performed (Supplementary Fig. 2b,c). This showed that nasal IFNs were consistently upregulated in infected participants, but other mediators (e.g., the innate cell chemoattracts, CCL3 and CCL4) were more heterogenous (Supplementary Fig. 2b). Peak mediator fold-changes from baseline were typically lower in the blood, but with a broader range of upregulated cytokines, including proinflammatory TNF-α, IL-6 and IL-18, and anti-inflammatory IL-10 that were not induced in nasal fluid (Supplementary Fig. 2c). Thus, the mucosal and systemic inflammatory mediator responses during mild SARS-CoV-2 infection were dominated by IFNs but distinct in timing and breadth, with a complex relationship that challenges the assumption that the circulating immune response is unidirectionally driven by, or a simple reflection of, inflammation at the site of infection.

### Polyclonal T cell responses peak at day 10 and 14 post-infection

Early cytokine and chemokine responses coordinate the recruitment, activation and expansion of immune cells. To assess the kinetics and magnitude of cell-mediated responses, polyclonal T cells were measured by flow cytometry using freshly isolated PBMC samples. CD38 and Ki67 were used to determine CD4^+^ and CD8^+^ T cell activation and proliferation (gated on single, live, CD3^+^ lymphocytes; gating strategy Supplementary Fig. 3a) (Fig. 3a). Participants who received a SARS-CoV-2 vaccine or were infected in the community before day 28 were excluded from this timepoint (see Supplementary Table 1). In infected participants, there was a significant increase in the frequency of CD38^+^Ki67^+^ CD4^+^ T cells, which peaked at day 10; while CD38^+^Ki67^+^ CD8^+^ T cells were elevated over a lengthier period; increasing from day 7 and peaking at day 14 (Fig. 3b,c). There was no increase in the frequency of activated, proliferating T cells over the course of infection in uninfected participants (Fig. 3b,c).

**Figure 3:**
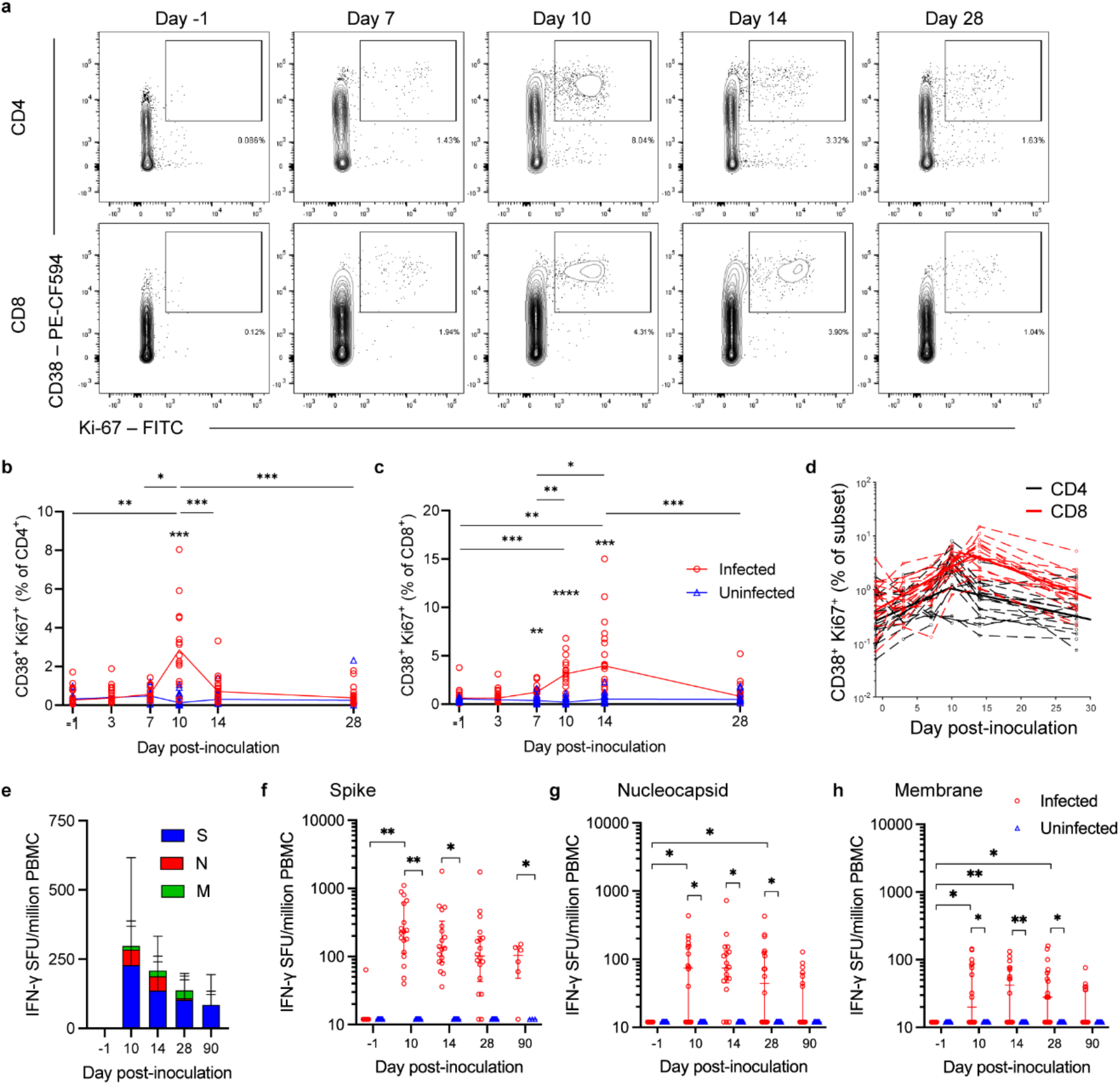
Activated and proliferating CD4^+^ and CD8^+^ T Cells display divergent kinetics post-infection. Polyclonal T cells in whole blood were co-stained with anti-CD4, CD8, Ki-67 and CD38. (A) Representative plots show CD38^+^Ki67^+^ CD4^+^ and CD8^+^ T cells (single, live, CD3^+^ lymphocytes; gating strategy shown in Supp Figure 1a). (B) CD4^+^ and (C) CD8^+^ T cell responses in the infected (red; n=18) and uninfected (blue; n=16) groups after inoculation are shown as median and individual points alongside (D) the piecewise linear regression model. (E) ELISpot IFN-y spot forming units (SFU) per million PBMC are shown in response to spike (S), nucleocapsid protein (N) and membrane (M) protein before and after inoculation in the infected group as stacked graphs (median and IQR). ELISpot responses in infected and uninfected groups against (F) S, (G) N and (H) M proteins are shown as median and IQR. Participants vaccinated and infected in the community were excluded from day 28 and day 90 data (see Supplementary Table 1 for details). Two-way ANOVA mixed-effects models with Geisser-Greenhouse correction for multiple testing was used to show significance between timepoints and groups. (*P* values *<0.05, **<0.01, ***<0.001, ****<0.0001).

Piecewise linear regression modelling showed the activated/proliferating CD8^+^ T cell response peaked later than the CD4^+^ T cell response, with the frequency of responding CD8^+^ T cells significantly higher by both AUC and peak size (Fig. 3d, Table 2). The growth and decay rates of activated and proliferating cells were not significantly different between CD4^+^ and CD8^+^ T cells (Table 2). All CD38^+^Ki67^+^ T cell frequencies returned to baseline levels by day 28.

**Table 2:**
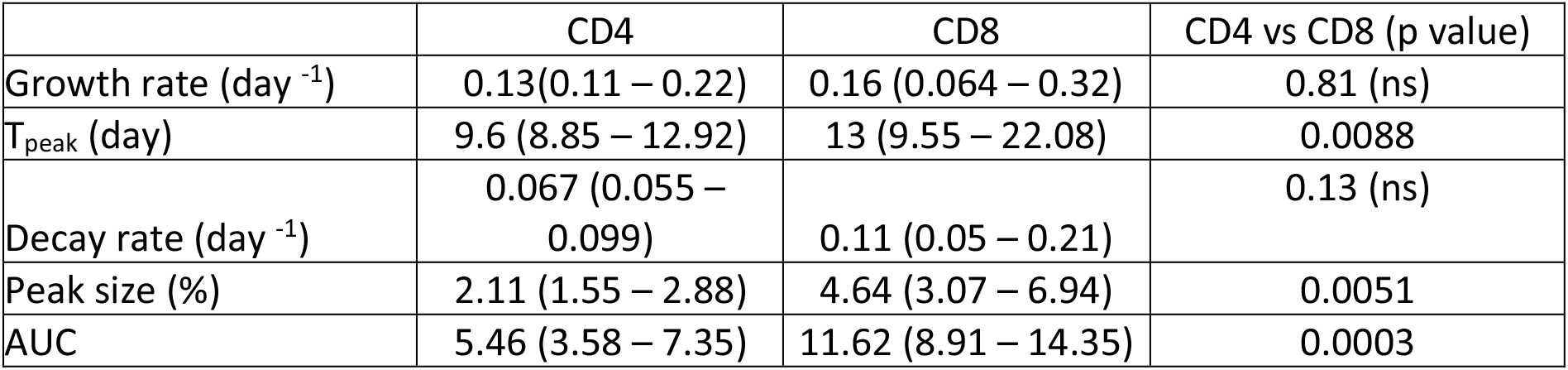
Modelled CD4 and CD8 T cell CD38^+^Ki67^+^ response parameters. Mean and 95% CI.

In contrast to Ki67, PD-1 co-expression on CD38^+^ T cells enabled earlier detection of significantly increased CD4^+^ T cell activation from day 7 to 14 post-infection and CD8^+^ T cells at days 7 and 10 (Supplementary Fig. 3b), suggesting that both CD4^+^ and CD8^+^ T cells are activated over a protracted period despite the shorter duration of CD4^+^ T cell proliferation. However, despite the expanded populations of activated cells, in the overall polyclonal population there was no increase in the frequency of PD-1^+^ or PD-1^+^LAG-3^+^ T cells and therefore no evidence of T cell exhaustion (Supplementary Fig. 3b-d). An increase in the frequency of CD4^+^ T cells expressing CXCR3 at day 10 post-inoculation in infected participants suggested a largely Th1-biased response (Supplementary Fig. 3e,f). CD4^+^ FoxP3^+^CD25^+^ Treg cells and CD45RA^-^CXCR5^+^ Tfh cells (cTfh) were gated and assessed for ICOS and PD-1 expression (Supplementary Fig. 3g). In the infected group, there was an increase in ICOS expressing Treg and cTfh cell frequency, and cTfh cells upregulated both PD-1 and ICOS indicating activation of these subsets (Supplementary Fig. 3h).

### Antigen-specific T cells are induced by day 10 post-inoculation

To quantify SARS-CoV-2 antigen-specific T cell responses, IFN-γ T cell ELISpots were performed on freshly isolated PBMC samples stimulated using overlapping peptides from spike (S) protein, nucleocapsid protein (NP) and membrane (M) protein. In the acute phase of infection (and excluding participants vaccinated and infected in the community before day 28 and day 90; Supplementary Table 1), the highest proportion of IFN-γ producing antigen-specific T cells were S protein-specific (Fig. 3e), with significant induction of S-specific T cells by day 10 post-inoculation compared with baseline (day -1) in infected participants only (Fig. 3f). S-specific responses then contracted but were still elevated at day 28 and 90 post-inoculation. NP and M specific T cell responses were also induced at day 10 and remained elevated until day 28 post-inoculation (Fig. 3g,h).

There were no detectable responses in the uninfected participants during the early post-inoculation period; however, induction of S-specific T cells was seen in the uninfected group from day 90 post-inoculation, coinciding with participants receiving SARS-CoV-2 vaccinations (ELISpot data at all follow-up timepoints with participants vaccinated and infected in the community included are shown in Supplementary Fig. 4a-c). No NP- or M-specific T cell responses were observed post-vaccination, but increased frequencies were seen in the uninfected group from day 90 onwards, following confirmed community SARS-CoV-2 infections (Supplementary Fig. 4).

### Immunodominant epitope-specific CD8^+^ T cells make up substantial proportions of the acute cell-mediated response during memory differentiation

To track and phenotype antigen-specific CD8^+^ T cells, fresh PBMC samples from 9 infected participants with appropriate HLA types were stained using MHC I-peptide pentamers based on previously described immunodominant epitopes (n=4 HLA-B*07:02 NP_105-113_ (SPRWYFYYL), n=5 HLA-A*02:01 S_269-277_ (YLQPRTFLL)^12^) (Fig. 4a). In infected participants, expanded populations of pentamer^+^ CD8^+^ T cells were detectable by day 14 post-inoculation, corresponding to the peak of polyclonal CD8^+^ T cell activation and proliferation (Fig. 4b). B*07:02 restricted NP-specific peak responses were immunodominant over A*02:01 restricted S-specific responses (median 2.045% B*07:02 NP vs median 0.84% A*02:01 S; ns by t-test). Antigen-specific CD8^+^ T cells remained elevated at day 28 post-infection but contracted by day 90, except in two B*07:02^+^ individuals with the highest peak responses (Fig. 4b).

**Figure 4:**
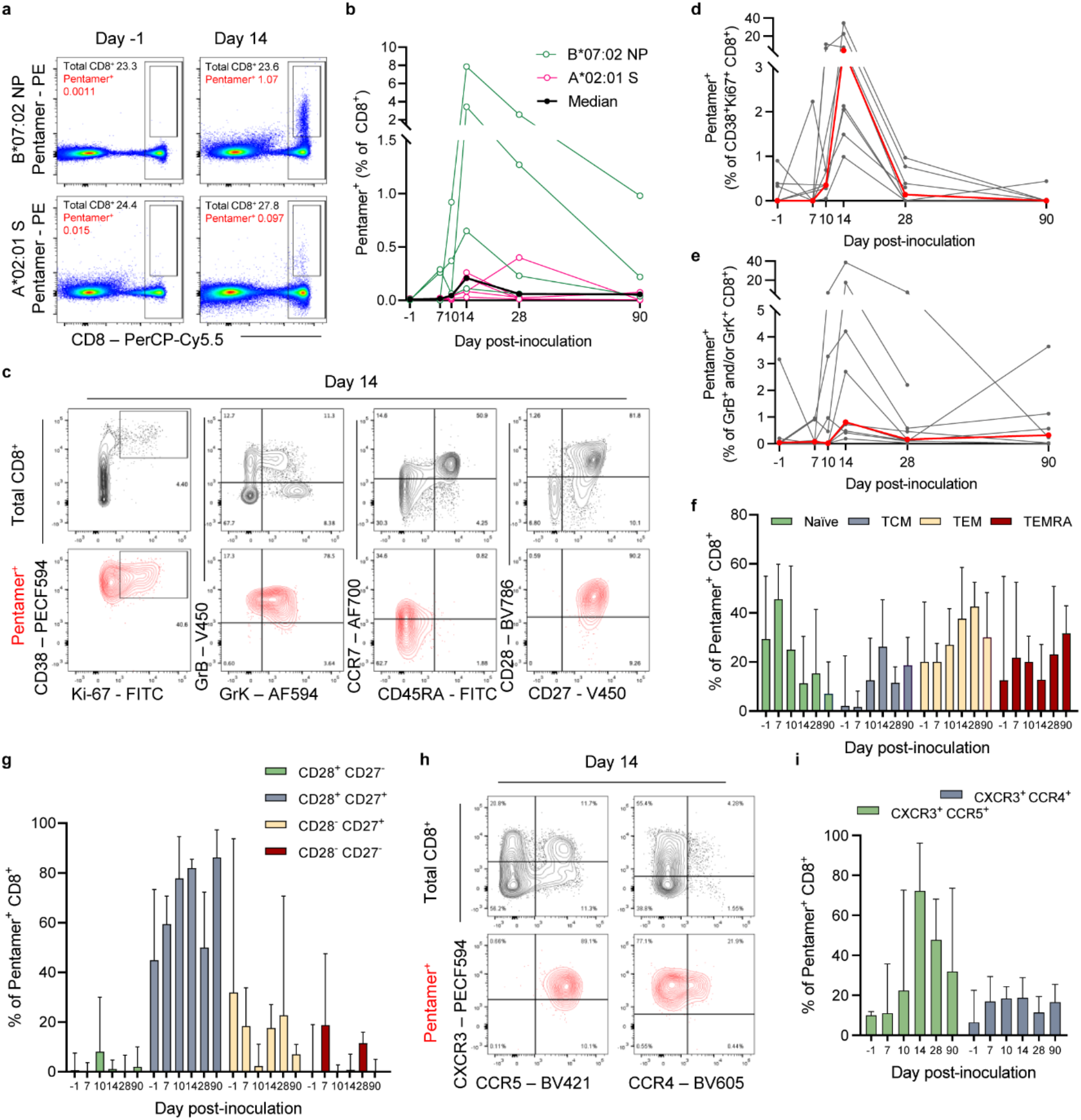
Antigen-specific CD8+ T cells are acutely activated, displaying central and effector memory phenotypes. (A) Representative plots show class I pentamer (PE conjugated B7-NP_105-113_ [SPRWYFYYL] and A2-S_269-277_ [YLQPRTFLL]) cells co-stained with anti-CD8. (B) The frequency of pentamer^+^ CD8^+^ T cells in the infected group with either B7-NP_105-113_ (green) or A2-S_269-277_ (red) and the median (black) are shown (total n=9). (C) Representative plots show the phenotype of total (black) and pentamer^+^ (red) CD8^+^ T cells. The proportion pentamer^+^ cells contributing to the (D) CD38^+^Ki67^+^ and (E) Granzyme (Gr) B and/or K-expressing CD8+ T cell populations are shown with individual data in black and medians in red. (F) Memory phenotype according to CCR7 and CD45RA expression and (G) CD27 and CD28 expression are shown as median and IQR. (H) Representative plots of CXCR3, CCR5 and CCR4 expression on total (black) and pentamer^+^ (red) CD8^+^ populations are shown (H), with (I) medians and IQR graphed. Participants vaccinated and infected in the community are excluded from day 28 and day 90 data (see Supplementary Table 1 for details). Two-way ANOVA mixed-effects models with Geisser-Greenhouse correction for multiple testing were used to test significance (all ns).

Phenotypic analysis showed that at the day 14 peak, large proportions of the activated/proliferating CD8^+^ T cell population were specific for single epitopes, with 4.05% (CI 1.49%-22.6%) of the total population of CD38^+^Ki76^+^ CD8 T cells labelled by the B*07:02-NP pentamer (median 15.07% (CI 4.05%-34.4%) and 2.05% (CI 0.99%-5.37%) by A*02:01-S (Figure 4c & 4d). Pentamer^+^ CD8 T cells also upregulated the cytotoxicity markers Granzyme (Gr) B and GrK, peaking at day 14, with median 0.81% of GrB and/or GrK expressing T cells being pentamer^+^ (Figure 4e).

Pentamer^+^ CD8^+^ T cells developed primarily effector memory (CCR7^-^CD45RA^-^) and to a lesser extent central memory (CCR7^+^CD45RA^-^) phenotypes at day 14, with decreasing frequencies of T cells with a naïve phenotype (CCR7^+^CD45RA^+^) after infection (Fig. 4f). Overall, the phenotype was indicative of an early memory response with only moderately increasing frequencies of TEMRA cells (CCR7^-^CD45RA^+^, more indicative of late or terminal differentiation) up to day 90 post-inoculation (Fig. 4f). Moreover, the majority of pentamer^+^ CD8^+^ T cells showed a less differentiated CD28^+^CD27^+^ phenotype at the peak with few cells downregulating CD28 and/or CD27 at day 14 and 28 (Fig. 4g). Acutely responding pentamer^+^ CD8^+^ T cells also displayed a CXCR3^+^CCR5^+^ phenotype, with low frequencies of CCR4^+^ T cells suggesting the upregulation of T cell homing capacity to sites of inflammation and the airway (Fig. 4h,i).

### Mucosal antibodies increase more rapidly than systemic responses but begin to wane early during convalescence

Daily nasal and plasma samples were analysed for virus-specific IgG, IgA, and IgM concentration against the SARS-CoV-2 Spike protein. Both nasal and systemic IgG, IgA and IgM responses began to appear around day 10 post-inoculation (Fig. 5a,b). Excluding participants who were vaccinated or infected in the community up to day 90 (Supplementary Table 1), levels remained low in the uninfected group (Fig. 5a,b). At day 28 plasma IgG and IgA levels were continuing to increase in the infected group but nasal titres plateaued between days 14 and 28 (Fig. 5a,b).

**Figure 5:**
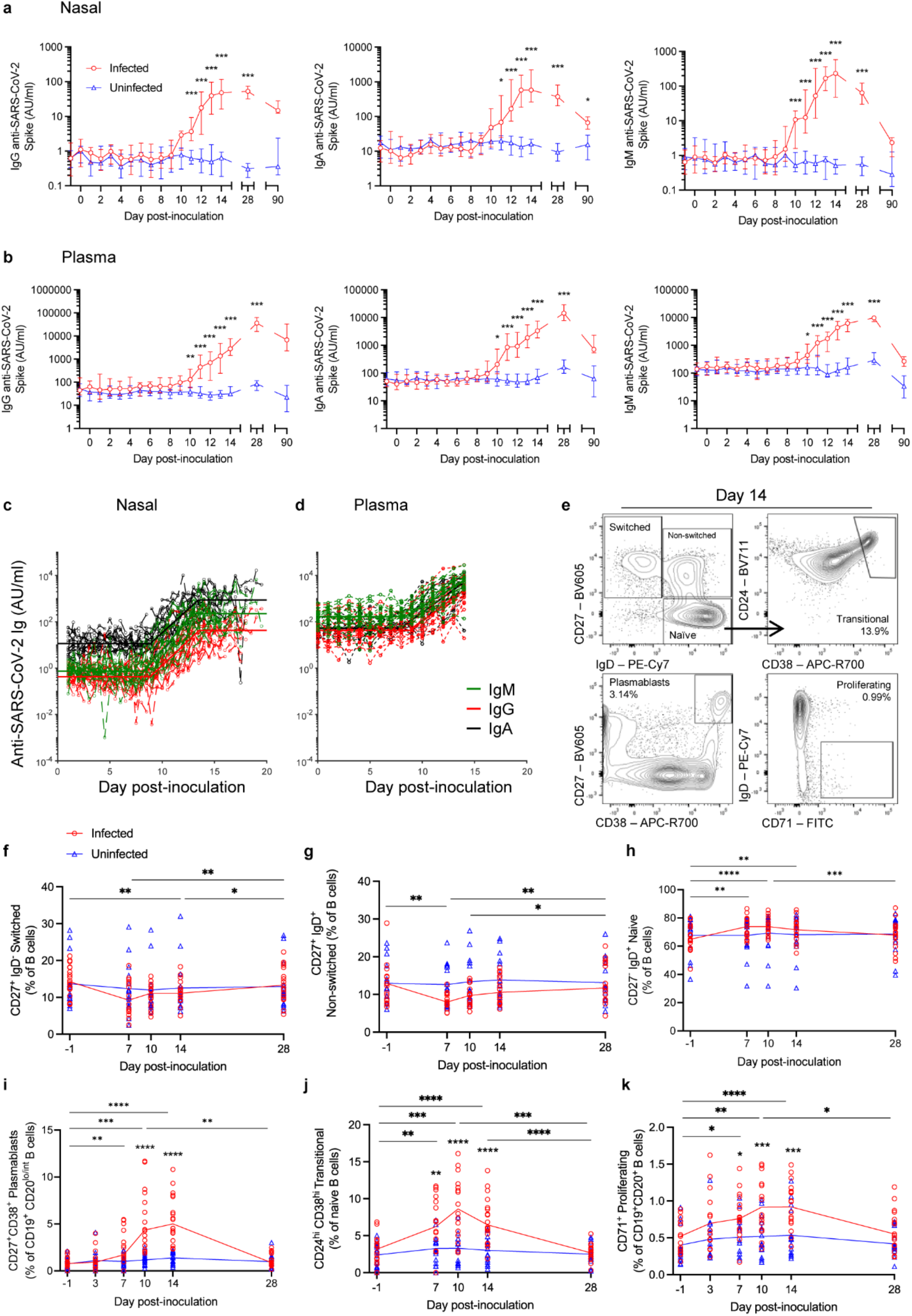
Mucosal and systemic antibody responses both appear from day 10 post-inoculation but plateau and wane more rapidly in the nose. Anti-spike antibody responses were measured by MesoScale Discovery and shown for each isotype (IgG, IgA and IgM) in (A) the nose and (B) plasma in infected (n=18) and uninfected (n=16) groups. Participants vaccinated and infected in the community were excluded from day 28 and day 90 data (see Supplementary Table 1 for details). The modelled antibody response in (C) the nose and (D) plasma are shown. (E) Representative plots show the gating strategy for CD19^+^ B cell phenotypes described by CD27 and IgD expression (switched, non-switched and naive), plasmablast (CD27^+^CD38^+^) transitional (CD24^hi^CD38^hi^), and proliferating IgD^-^ B cells (CD71^+^). (F-K) Frequencies of each subset in infected and uninfected groups after inoculation are shown as median and individual points. Significance between infected and uninfected groups was tested by multiple Mann-Whitney tests with Holm-Šídák’s correction for testing of multiple timepoints (antibody). Two-way ANOVA mixed-effects models with Geisser-Greenhouse correction for multiple testing was used to show significance between timepoints and groups (B cells). (*P* values *<0.05, **<0.01, ***<0.001, ****<0.0001).

Piecewise linear modelling was again employed to more precisely determine the kinetics of the anti-SARS-CoV-2 Spike antigen antibody response (Fig. 5c,d). All three antibody isotypes, both in the nose and the plasma, were induced at a similar time (T_activation_ between day 8 and 9) (Table 3). The peak size and AUC of all three isotypes were higher in the plasma than in the nose (Table 3). However, the mucosal antibody response increased more rapidly, with growth rates for all isotypes higher in the nose than the plasma (Table 3).

**Table 3:** Modelled nasal and systemic antibody response parameters. Mean and 95% CI.

|  | IgG |  |  | IgA |  |  | IgM |  |  |
| --- | --- | --- | --- | --- | --- | --- | --- | --- | --- |
|  | Nasal | Plasma | Nasal vs plasma (p value) | Nasal | Plasma | Nasal vs plasma (p value) | Nasal | Plasma | Nasal vs plasma (p value) |
| T<sub>activation</sub> (day) | 8.85<br>(8.07 – 9.69) | 8.87<br>(7.08 – 11.11) | 0.97<br>(ns) | 9.03<br>(8.34 – 9.76) | 7.91<br>(6.4 – 9.61) | 0.03 | 8.25<br>(7.7 – 8.83) | 8.33<br>(6.97 – 9.94) | 0.86<br>(ns) |
| Growth rate (day<sup>-1</sup>) | 0.42<br>(0.37 – 0.48) | 0.34<br>(0.24 – 0.47) | 0.04 | 0.42<br>(0.37 – 0.47) | 0.33<br>(0.24 – 0.44) | 0.008 | 0.51<br>(0.45 – 0.58) | 0.27<br>(0.19 – 0.38) | 9.9x10<sup>-9</sup> |
| T<sub>peak</sub> (day) | 13.60<br>(12.93 – 14.3) | 16.83<br>(11.47 – 24.69) | 0.21<br>(ns) | 13.60<br>(12.91 – 14.3) | 13.68<br>(11.86 – 15.78) | 0.90<br>(ns) | 13.20<br>(12.6 – 13.83) | 14.64<br>(12.45 – 17.22) | 0.08<br>(ns) |
| Peak size (log10) | 2.4 (2.1 – 2.71) | 4.73<br>(4.43 – 5.05) | 4.92x10<sup>-13</sup> | 3.36<br>(3.12 – 3.61) | 4.19<br>(4.02 – 4.36) | 1.3x10<sup>-6</sup> | 2.76<br>(2.47 – 3.07) | 3.99<br>(3.87 – 4.11) | 3x10<sup>-9</sup> |
| AUC (log10) | 3.91<br>(3.58 – 4.23) | 6.39<br>(6.12 – 6.67) | 4.92x10<sup>-14</sup> | 4.53<br>(4.36 – 4.71) | 5.85<br>(5.69 – 6.02) | 2.2x10<sup>-13</sup> | 3.75<br>(3.54 – 3.95) | 5.63<br>(5.52 – 5.75) | 2.6x10<sup>-18</sup> |

Including all participants up to day 360 post-inoculation (including those vaccinated and infected in the community), there was a significant induction of virus-specific IgG in the nose and plasma of uninfected participants at day 180 onwards compared with baseline levels (Supplementary Fig. 5a,b). Plasma IgA was also significantly higher than baseline at day 180 in the uninfected group (Supplementary Fig. 5b). In the infected group, nasal and plasma IgG remained elevated at day 360; however, virus-specific nasal IgA and IgM (and plasma IgM) began to decline after day 28 and was not significantly boosted thereafter (Supplementary Fig. 5a). Plasma IgA was significantly boosted at day 270 and day 360 after an initial decline at day 90. Grouping data by the timing of first vaccination dose (early before day 90 or later after day 90) (Supplementary Fig. 5c,d), vaccine-induced antibody increases in both the infected and uninfected groups were evident irrespective of timing of vaccination.

Phenotypic analysis of B cells using flow cytometry at the same timepoints as the T cell analysis (gating strategy Fig. 5e) showed a decrease in the frequency of switched and non-switched B cells and a reciprocal increase in the frequency of naïve B cell populations at day 7 and 10 after inoculation compared with day -1 in the infected group (Fig. 5F-H). Plasmablasts were induced at day 10 and frequencies remained elevated over day -1 through to day 14 (Fig. 5i). Transitional and proliferating B cell populations were also induced at day 7, 10 and 14 (Fig. 5j,k). There were no changes in B cell frequencies in the uninfected group. Thus, local antibody response following primary infection was induced more rapidly but with an earlier and lower peak than antibodies in the circulation. While both vaccination and community infection were associated with nasal and plasma IgG and plasma IgA increases, a substantial increase in nasal IgA and IgM was not detected after vaccination or community infection.

### CD8^+^ T cell and antibody responses are associated with viral load decline

Using the quantitative and kinetic parameters estimated by mathematical modelling of the VL and immune measures, we further analysed the relationship between them to infer potential drivers of the immune response and reciprocal viral suppression. Earlier VL detection time and a more rapid VL growth rate strongly correlated with early nasal IFN activation and peak time (Fig. 6a). In addition, earlier IFN activation and peak times correlated with greater VL AUC. Similarly, the VL growth rate and nasal IL-29 growth rate were highly correlated (Fig. 6b). However, there was no significant relationship between the magnitude of nasal IFNs and VL decay rates (not shown). No significant correlations were seen with plasma IFN responses (not shown). Together, the strong correlations in kinetics suggest that VL drives the local interferon response, but higher mucosal interferon levels did not appear to directly expedite viral clearance.

**Figure 6:**
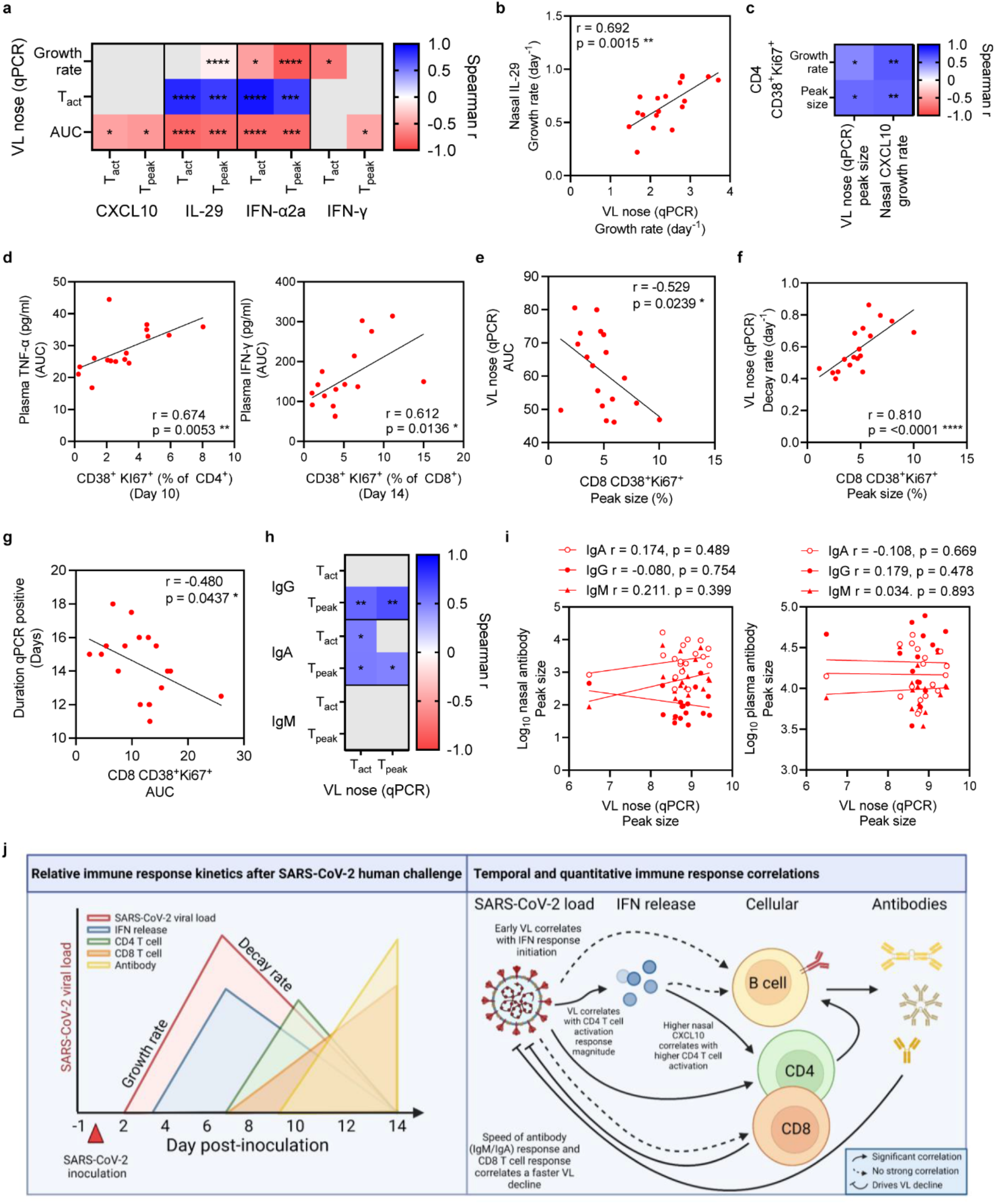
Activated and proliferating CD8^+^ T cell and early mucosal IgM and IgA responses correlate with viral load decline. Piecewise linear regression models were fitted to viral load (VL) and immune response data. Heat maps and XY plots show the correlation between VL and immunological parameters. (A) Heat map shows Spearman correlations between VL parameters and T_activation_ and T_peak_ of nasal CXCL10, IL-29, IFN-α2a and IFN-y. (B) Correlation of growth rate of nasal IL-29 and VL is shown. (C) Correlations between CD4^+^ T cell activation and proliferation (growth rate and peak size) and VL peak size and nasal CXCL10 growth rate. (D) The relationship between CD4^+^ and CD8^+^ T cell responses and plasma TNF-α (left) and IFN-y (right) AUC. (E) Corelations between peak CD8^+^ T cell activation and proliferation and VL AUC, (F) decay rate and (G) the duration of qPCR positivity. (H) The relationship between nasal antibody T_activation_ and T_peak_ and VL T_activation_ and T_peak_. (I) The relationship between nasal/plasma antibody and VL peak size. (J) Non-significant correlations are shown as grey in the heatmaps. (J) A schematic summarises the statistical relationships between VL and immune response, created with BioRender.com. Spearman correlation. (*P* values *<0.05, **<0.01, ***<0.001, ****<0.0001).

Similarly, VL peak size correlated significantly with the growth rate and peak size of CD38^+^Ki67^+^ CD4^+^ T cell frequency suggesting a close relationship between VL and the speed and magnitude of the CD4^+^ T cell response (Fig. 6c). In addition, a higher nasal CXCL10 growth rate was associated with higher CD38^+^Ki67^+^ CD4^+^ T cell growth rate and peak size (Fig. 6c), implying coordination of the interferon-stimulated CXCL10 expression with activated CXCR3^+^ T cells generated during acute infection. Thus, both VL and associated interferon-stimulated responses correlate with CD4^+^ T cell activation and proliferation. Furthermore, a higher frequency of CD38^+^Ki67^+^ CD4^+^ and CD8^+^ T cells was associated with higher plasma IFN-γ and TNF-α concentration AUCs, consistent with T cells as a source of these cytokines in the periphery (Fig. 6d).

In contrast, the relationship between the CD8^+^ T cell response and VL differed, with no correlation between VL and timing of activated CD8^+^ T cell response but higher CD38^+^Ki67^+^ CD8^+^ T cell peak size negatively correlating with VL AUC (Fig. 6e). Moreover, the peak frequency of activated/proliferating peripheral CD8^+^ T cells correlated with a higher VL decay rate (Fig. 6f) and a shorter duration of infection by PCR positivity in days (Fig. 6g). There was no significant correlation between CD4^+^ T cell or B cell responses and VL decay rate (Supplementary Fig. 6a). Together, these data strongly support the role of CD8^+^ T cell activation and proliferation as a dynamic marker and likely effector of viral clearance during primary human SARS-CoV-2 infection.

As for IFN and CD4^+^ T cell responses, earlier VL T_activation_ and T_peak_ were associated with earlier nasal IgA activation time and peak time, and the same was true for nasal IgG (Fig. 6h). There was no association between VL peak size and antibody peak size, suggesting that while VL triggers the antibody response, it does not determine the magnitude (Fig. 6i). Significant correlations were evident between VL decay rates and the growth rate of IgM (r=0.58, p=0.012) and IgA (r=0.57, p=0.013), but not IgG. The appearance of virus-specific antibodies was closely aligned with the end of replication-competent virus shedding, with similar timing between a 4-fold rise in nasal IgA and FFA-negativity on average. Together, these observations suggest that not only CD8^+^ T cells but also *de novo* mucosal IgM and IgA responses are generated in a timeframe relevant to acute infection and may contribute to termination of infectivity.

The activation time of CXCL10 correlated positively with the activation (r=0.5; p=0.033) and peak time (r=0.64; p=0.004) of the plasmablast response, suggesting the B cell response was also initiated in accordance with the start of the IFN-stimulated response. A higher CD38^+^Ki67^+^ CD4^+^ T cell peak size was also associated with earlier nasal antibody activation time (IgA r=-0.53, p=0.023; IgG r=-0.47, p=0.046; IgM r=-0.5, p=0.035) and peak size (IgA r=0.48, p=0.04; IgM r=0.56, p=0.016). In view of the temporal sequence of CD4^+^ T cell and B cell activation, this further supports the importance of CD4^+^ T cell help when antigen-specific B cells are most plentiful to promote development of the subsequent antibody response. However, in the primary infection setting, there was no significant correlation between low-frequency CD4^+^ cTfh cell and B cell responses or the resulting antibody levels measured at day 28 (Supplementary Fig. 6b). The relationships measured between viral and immune responses are summarised in a schematic (Fig. 6j).

## Discussion

Experimental infection of volunteers can reveal the exact timing and sequence of events during each stage of infection and viral clearance, and the relation of these events to prior immune status. The granularity and depth of analysis allows precise modelling of human immune responses, beyond that which is possible in any other setting. The situation of the studies that we now describe is especially unusual in that our volunteers, along with the rest of the population, were entirely naïve to this novel virus and had not yet been vaccinated at the start of the experiments. Here we have provided a detailed analysis of the sequential viral, inflammatory and immune events in both nasal mucosa and circulation that permits their quantitative and temporal relationships to be revealed.

In participants who showed sustained infection, mucosal and systemic soluble mediator responses were dominated by IFNs with distinct kinetics and few correlations between the nasal and systemic compartments. While these data indicate that virus in the nose triggers IFN release within the first 24 hours, some IFN-related responses appeared earlier in the blood than at the site of infection, likely due to rapid trafficking of innate cells in and out of these compartments. Indeed, single cell RNA sequencing of samples from these same participants showed this to be mainly derived from monocytes and DCs^13^, with further mechanistic explanation for their rapid response provided by epigenetic analyses indicating differential accessibility of ISGs in these different cell types^14^. Together these data challenge the expectation that responses to respiratory viral infection invariably start in the mucosa to be followed by events seen in the circulation. Thus, diagnostic markers in the blood may in fact be a more sensitive early post-viral exposure test than were previously^14^.

Innate IFNs are a critical component of the anti-viral immune response; reduced type I IFN receptor expression is associated with severe COVID-19 disease, and polymorphisms in IFN, *IFNAR* and several ISGs are associated with COVID-19 severity. Furthermore, autoantibodies that neutralise IFNs may stall this response and are similarly associated with severity and imbalanced IFN responses are observed in severe disease^10, 15, 16^. However, while the timing and magnitude of VL and IFN levels were well correlated, there was no evidence that IFNs at these physiological levels directly modulated the magnitude of virus replication or led to the decline of viral load. This suggests that immediate IFN activity may have less impact in the context of mild infection, perhaps due to SARS-CoV-2-mediated IFN-evasion mechanisms^17^, although this does not preclude a role for IFNs in the context of severe disease.

We previously reported, using the same assay, substantially higher IL-6 levels at 10-100 pg/ml in plasma from hospitalised patients, relative to peak medians below 1 pg/ml in these mild experimental infections^18^. Interestingly, plasma IFN-γ levels appear more similar between these mild infections (peak median 10-20 pg/ml) and hospitalised patients (median 10-100 pg/ml), again using the same assays, suggesting that a relatively normal IFN response may be overlaid by an exaggerated inflammatory response in severe COVID-19. Our data also supports the lack of a strong association between viral load and symptom severity noted elsewhere^19^, while indicating an association between peripheral immune responses and the mild symptoms experienced by our volunteers.

Earlier studies have indicated that a rapid and robust T cell response is critical in determining the outcome of SARS-CoV-2 infection^20, 21^. Here, we showed T cell responses as early as day 7, with peak T cell activation and proliferation at day 10 and day 14 and an antigen-specific T cell response against Spike dominant in most infected participants. The kinetics of T cell activation and proliferation were distinct between CD4^+^ and CD8^+^ T cell populations, the CD8^+^ T cell response being earlier and more prolonged. However, compared with other respiratory virus challenge studies, which are conducted in healthy adults who have had multiple previous infections, the time to peak of CD38^+^Ki67^+^ CD8^+^ T cell expression was delayed, contrasting with peaks at day 10 and day 7 after RSV and influenza virus infection respectively^8, 22^. Field studies have shown that multiple SARS-CoV-2 exposures or vaccinations shift the T cell memory population to a more differentiated and TEMRA phenotype^23^. However, the early memory phenotype of antigen specific CD8^+^ T cells and the relatively late peak were consistent with these being newly primed T cell responses as opposed to reactivation of memory cells.

Despite our understanding that these primary T cells may be less functionally capable than their memory counterparts, our analysis implicates these CD8^+^ T cell responses as a major factor in viral clearance. Spike and nucleocapsid protein-specific CD8^+^ T cells upregulated cytotoxic molecules, suggesting a potential role of CD8^+^ T cell cytotoxicity in viral clearance. SARS-CoV-2 infection has been shown to be associated with a predominance of CD8^+^ T cell activation over CD4^+24^ but, in our analysis, the main driver of the CD8^+^ T cell response remains unclear as neither VL or IFN correlated with CD8^+^ T cell activation and proliferation. The uncoupling of CD8^+^ T cell and IFN responses has been seen in other studies ^24^ and may suggest involvement of pre-existing cross-reactive memory T cells detectable in some SARS-CoV-2 unexposed individuals and where IFN-mediated “signal 3” is less critical^25–27^. These may aid or modulate the T cell response and/or contribute directly to viral clearance in these participants. A robust CD8^+^ T cell response is therefore likely to be a major effector mechanism in self-limiting/mild infection and therefore interventions to enhance this, such as vaccines to generate T cells recognising more conserved epitopes for broadly-protective vaccines, should be prioritised.

While the CD8^+^ T cell response appeared to affect clearance of viral RNA, the timing of the end of replication-competent viral shedding (measured by FFA) was closely associated with antibody responses, especially those in the mucosa. Antibody responses in the blood and nose were largely synchronous in their initiation, from around day 9, but markedly diverged thereafter. Piecewise linear regression showed that the growth rate, peak and scale of antibody responses differed between the blood and nose. Similarly, anti-Spike antibodies in plasma continued to rise beyond day 14 while their mucosal counterparts had already started to fall, supporting the compartmentalisation of mucosal antibody responses we have previously reported after severe COVID-19^28^. The lack of boosting seen in nasal IgA and IgM following vaccination, in contrast to nasal IgG and plasma IgA, is in concordance with other studies of vaccination after infection^28^. However, our data show that boosting of mucosal antibodies may be also limited after recurrent infection, suggesting that boosting of local antibodies may be difficult to achieve.

The integration of VL with nasal and plasma IFN, T cell, B cell and antibody responses in this unique, serologically naïve cohort thus reveals a complex and intricate relationship between the different arms of the immune response and between parameters measured at the site of infection (the nose) and in the peripheral blood. With measurements of transmissible virus shed in the environment suggesting that emitted virus is most strongly correlated with nasal VL,^29^ these findings highlight the critically important role of rapid viral recognition and dynamic immune responses, particularly emphasising CD8^+^ T cells as likely effectors of viral clearance to limit both disease and transmission.

## Data Availability

All data produced in the present study are available upon reasonable request to the authors.

## Acknowledgements

We thank the study participants for their time and commitment. The authors gratefully acknowledge support from the UK Vaccine Taskforce of the Department of Business, Energy and Industrial Strategy of Her Majesty’s Government (BEIS) and the Wellcome Trust (grant no. 224530/Z/21/Z). We thank Oxford Immunotec, Oxford, U.K. for performing the ELISpot assay. CC and PO are supported by the NIHR Imperial Biomedical Research Centre (BRC) award to Imperial College Healthcare NHS Trust and Imperial College London. PO is supported by an NIHR Senior Investigator Award (NIHR201385) and UKRI MRC CIC Award (MR/V028448/1). MPD and AR are supported by Australian NHMRC grants 1149990 and 1173027, and MRFF award 2005544. The views expressed are those of the authors and not necessarily those of the NHS, the NIHR, DHSC or BEIS.

## Author contributions

H.R.W., R.S.T. processed samples, acquired data, analysed and interpretated data and wrote the manuscript. A.R. analysed and interpretated data and wrote the manuscript. J.K.S., R.M., S.A., A.M.C., J.X., L.P., N.L. processed samples and acquired data. M.K.S., B.M.C. analysed and interpretated data. B.K., M.K., A.M., A.C. oversaw sample collection and provided clinical data. M.P.D., P.O. interpreted data and wrote the manuscript. C.C. conceived the study, set up the clinical study, directed the study, interpreted data and wrote the manuscript.

## Methods

### Study design, ethical approval and participants

The study was conducted in accordance with the protocol; the consensus ethical principles derived from international guidelines, including the Declaration of Helsinki and Council for International Organizations of Medical Sciences International Ethical Guidelines; applicable ICH Good Clinical Practice guidelines; and applicable laws and regulations. The study design, public consultation, and ethical approvals for the study have been detailed previously ^9^.

Briefly, the screening protocol and main study were approved by the UK Health Research Authority’s Ad Hoc Specialist Ethics Committee (references 20/UK/2001 and 20/UK/0002) and registered with ClinicalTrials.gov (identifier NCT04865237). Volunteers were recruited from respondents to the study advertisements and were healthy adults aged 18-30 years with no previous SARS-CoV-2 infection or vaccination. The study was conducted in a high-containment clinical trials unit at the Royal Free London NHS Foundation Trust where participants were housed in single-occupancy, negative pressure side rooms. Participants were inoculated intranasally by pipette with 10 TCID50 of wild-type SARS-CoV-2 (20A clade of the B.1 lineage possessing the D614G mutation; GenBank accession number OM294022) between both nostrils (100 µl each). Safety was assessed with daily blood tests, spirometry, electrocardiograms, clinical assessments (vital signs, symptom diaries and clinical examination) and CT scan of chest on day 5 (all participants) and day 10 (infected participants only).

### Sample and data collection

Venous blood plasma (EDTA) and nasosorption samples ^30^ were collected daily for the measurement of peripheral and mucosal mediators and antibodies, respectively. Venous blood (lithium heparin) samples were collected at day -1 (baseline), 3, 7, 10, 14, 28, 90, 180, 270 and 360 for PBMC isolation. Nasosorption samples were eluted in 330µl of immunoassay buffer AB-33k (Millipore) containing 1% Triton-X100 (v/v) (Sigma).

Virological assessments of infections were based on 12-hourly mid-turbinate and throat flocked swabs placed in 3ml of viral transport medium (BSV-VTM-001, Bio-Serv) that was aliquoted and stored at −80 °C. Aliquots were analysed by RT-PCR and quantitative culture by FFA as previously described ^9^. Symptom diaries were self-completed three times daily from the day -1 (baseline) to day 14 after inoculation. A total of 19 symptoms covering upper respiratory, lower respiratory and systemic symptoms were scored on a scale of 0–3 (0=no symptoms, 1=mild, 2=moderate and 3=severe). Viral load and symptom data was reported previously ^9^.

### Quantification of soluble mediators and antibody by MesoScale Discovery

A panel of 35 cytokine and chemokine immune mediators were measured by MesoScale Discovery (MSD) multiplex immunoassays, consisting of CCL2, CCL3, CCL4, CCL13, CCL22, CXCL10, Eotaxin, Eotaxin-3, GM-CSF, IFNα2a, IFNβ, IFN-γ, IFN-λ (IL-29) IL-1α, IL-1β, IL-2, IL-4, IL-5, IL-6, IL-7, IL-8 (CXCL8), IL-10, IL-12p40, IL-12p70, IL-13, IL-15, IL-16, IL-17A, IL-18, IL-33, TARC, TNFα, TNFβ, TSLP and VEGF-A per manufacturer’s instructions. Unquantifiable samples were given a value of the lower limit of detection, denoted on plots as a dashed line. SARS-CoV-2 Spike antibody titres were similarly quantified using MSD multiplex immunoassay Coronavirus panel 2. Antibody panels were developed using anti-human IgG, IgM, or IgA and binding titres given as arbitrary units per milliliter (AU/ml) based on a kit-provided human plasma standard curve.

### ELISpot

*Ex vivo* ELISpot was carried out by Oxford Immunotec as follows. Peripheral blood mononuclear cells (PBMCs) were isolated from the whole blood samples by density gradient centrifugation method (Ficoll-Paque®). The addition of T-Cell Xtend® reagent (Oxford Immunotec Abingdon, UK) to extend peripheral blood mononuclear cell (PBMC) survival was used. The PBMCs were counted using flow cytometry to ensure an adequate number in all patients samples.

IFN-γ secreting T cells specific to specific antigen panels (Spike protein N-terminus, Spike protein C-terminus, Nucleocapsid protein and Membrane protein) based on the Wuhan-Hu-1 sequence (YP_009724390.1) were detected using the T-SPOT® Discovery test performed, according to the manufacturer’s protocol, at Oxford Immunotec within 32 h of venepuncture. The PBMCs were also incubated with a Nil control to identify the non-specific cell activation, and a positive control to ensure T cell reactivity was present. Antigen-specific T cell frequencies were reported as spot forming cells (SFC) per 250,000 PBMCs.

### Flow cytometry

PBMCs were isolated by density centrifugation using Histopaque 1077 (Sigma-Aldrich) according to the manufacturers protocol. Briefly, whole blood samples were diluted (1:1) in PBS and overlayered onto Histopaque, centrifuged for 30 minutes at 400xg. Isolated PBMCs were washed in PBS and used immediately. 1×10^6^ PBMC were stained with PE-conjugated class I pentamer A*02:01 YLQPRTFLL-PE (Spike) or B*07:02 SPRWYFYYL-PE (N) (Proimmune) for 10 min at room temperature. Cells were washed with PBS and stained with Zombie UV Fixable Viability Kit (Biolegend), followed by staining with antibodies against surface markers in FACS buffer (PBS containing 2% FCS and 2 mM EDTA). Cells were washed and either fixed with BD CellFIx (BDBiosciences) or fixed and permeabilised for intranuclear or intracellular staining using Foxp3/Transcription Factor Staining Buffer Set (ThermoFisher).

Antibodies for staining as follows; CD8 PerCP-Cy5.5 (eBioscience, ThermoFisher), PD-1 BV605, CD4 APC-H7, CD38 PE-CF594, Ki67 FITC, Granzyme B V450, CD45RA FITC, CD27 V450, CD28 BV786, CXCR3 PE-CF594 CXCR3 PerCP-Cy5.5, CCR6 BV786, FoxP3 PE-CF594, CD25 PE-Cy7, ICOS BV650, CD27 BV605, IgD PE-Cy7, CXCR5 BV711, CD19 BV650, CD20 APC-H7, CD38 APC-R700, CD71 FITC (all BDBiosciences), CD3 BV510, LAG-3 BV650, CCR7 AF700, CCR5 BV421, CCR4 BV605, CD24 BV711 (all Biolegend) and Granzyme K AF594 (Santa Cruz, Biotechnology). Cells were washed in FACS buffer or permeabilisation buffer and stored in FACS buffer until acquisition. Samples were analysed using a LSRFortessa (BDBiosciences) and FlowJo software (version 10.8.1). Remaining PBMC were cryopreserved in FBS (Sigma-Aldrich) with 10% DMSO in liquid nitrogen.

### Modelling

We used a piecewise model to estimate the activation time, peak time, growth rate and decay rate of various immune responses (see Figure 1B for the schematic of the model). The model of the immune response y for subject i at time y_!_ can be written as:

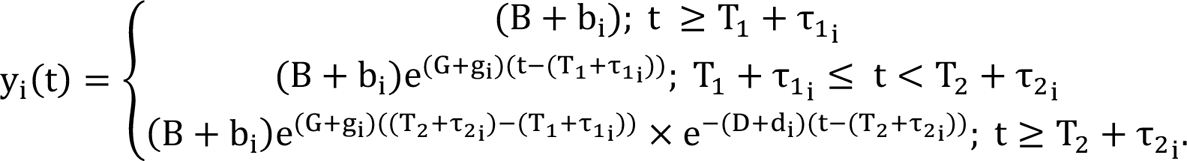

The model has 5 parameters; B, G, T_1_, D, and T_2_. For a period before T_1_, we assumed a constant baseline value B for the immune response (which is higher than or at the background level). After the activation time T_1_, the immune response will grow at a rate of G until T_2_. From T_2_, the immune response will decay at a rate of D. For each subject i, the parameters were taken from a normal distribution, with each parameter having its own mean (fixed effect). A diagonal random effect structure was used, where we assumed there was no correlation within the random effects. The model was fitted to the log-transformed data values, with a constant error model distributed around zero with a standard deviation σ.

For the viral levels, the initial level was below the level of detection, and values less than the limit of detection were censored in the fitting. We estimated a T_activation_ for virus (for comparison purposes) as the time when the VL is predicted to cross the level of detection (1000 copies). To account for the values less than the limit of detection, a censored mixed effect regression was used to fit the model. Model fitting was performed using MonolixR2019b.

### Statistics

Statistical analysis was performed using GraphPad Prism version 9.2 and R version 4.05. Longitudinal comparisons between infected and uninfected groups utilised multiple Mann-Whitney tests with Holm-Šídák’s correction for multiple testing (soluble mediators and antibody) and two-way ANOVA mixed-effects models with Geisser-Greenhouse correction for multiple testing. Post-hoc testing was carried out using Tukey’s Test analysis to account for multiple comparisons (T cells and B cells). Correlation analyses used Spearman. A p value of less than 0·05 was used to indicate significance.

## Supplementary Information

**Supplementary Figure 1:**
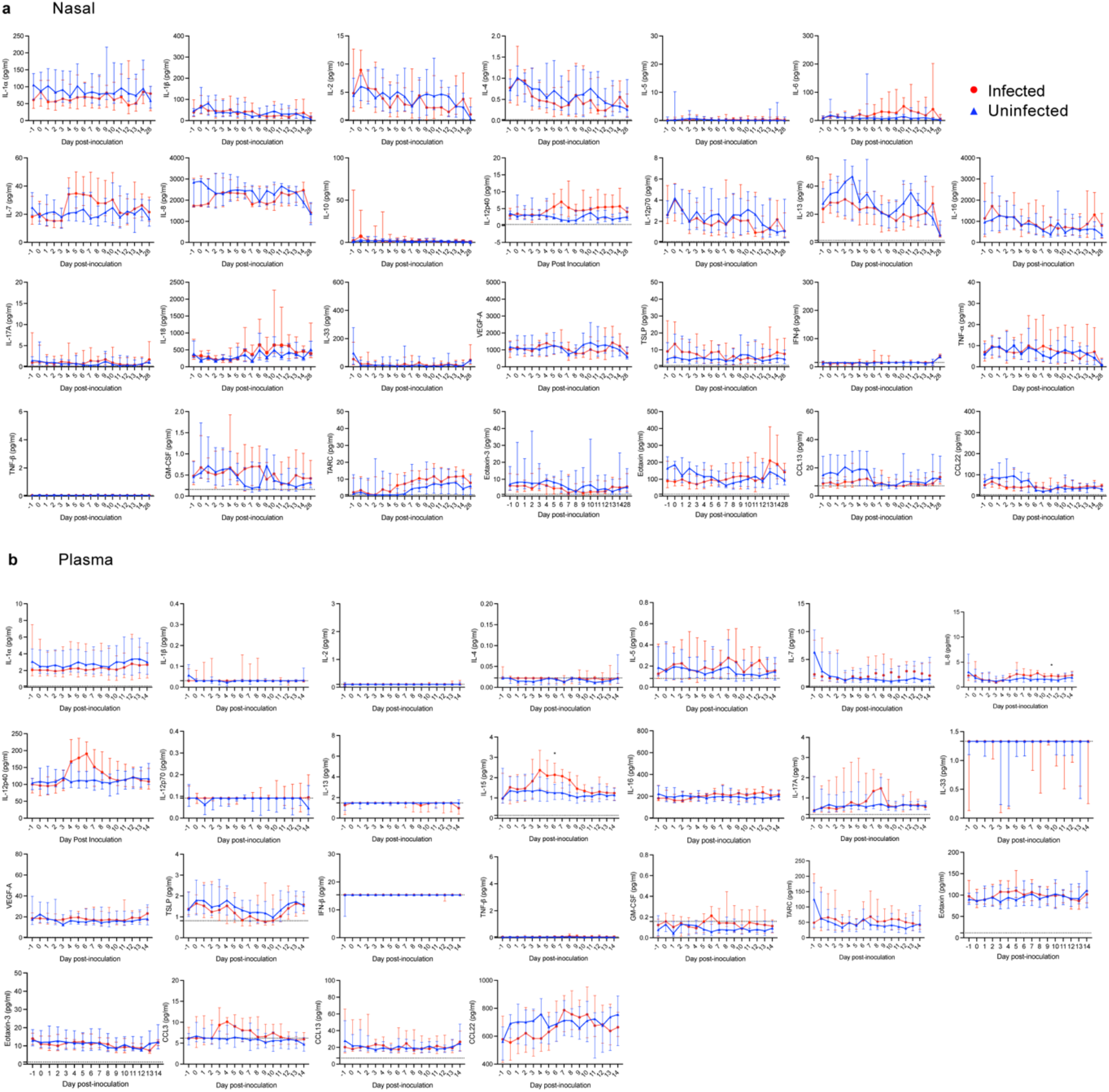
The local mucosal and systemic soluble mediator response following SARS-CoV-2 inoculation. Figures show the indicated soluble mediator level in (A) the nose and (B) plasma in the infected (n=18) and uninfected groups (n=16) after inoculation as median and IQR. Participants vaccinated and infected in the community are excluded from day 28 data (see Supplementary Table 1 for details). Significance between infected and uninfected groups was tested by multiple Mann-Whitney tests with Holm-Šídák’s correction for testing of multiple timepoints (adjusted P values *<0.05).

**Supplementary Figure 2:**
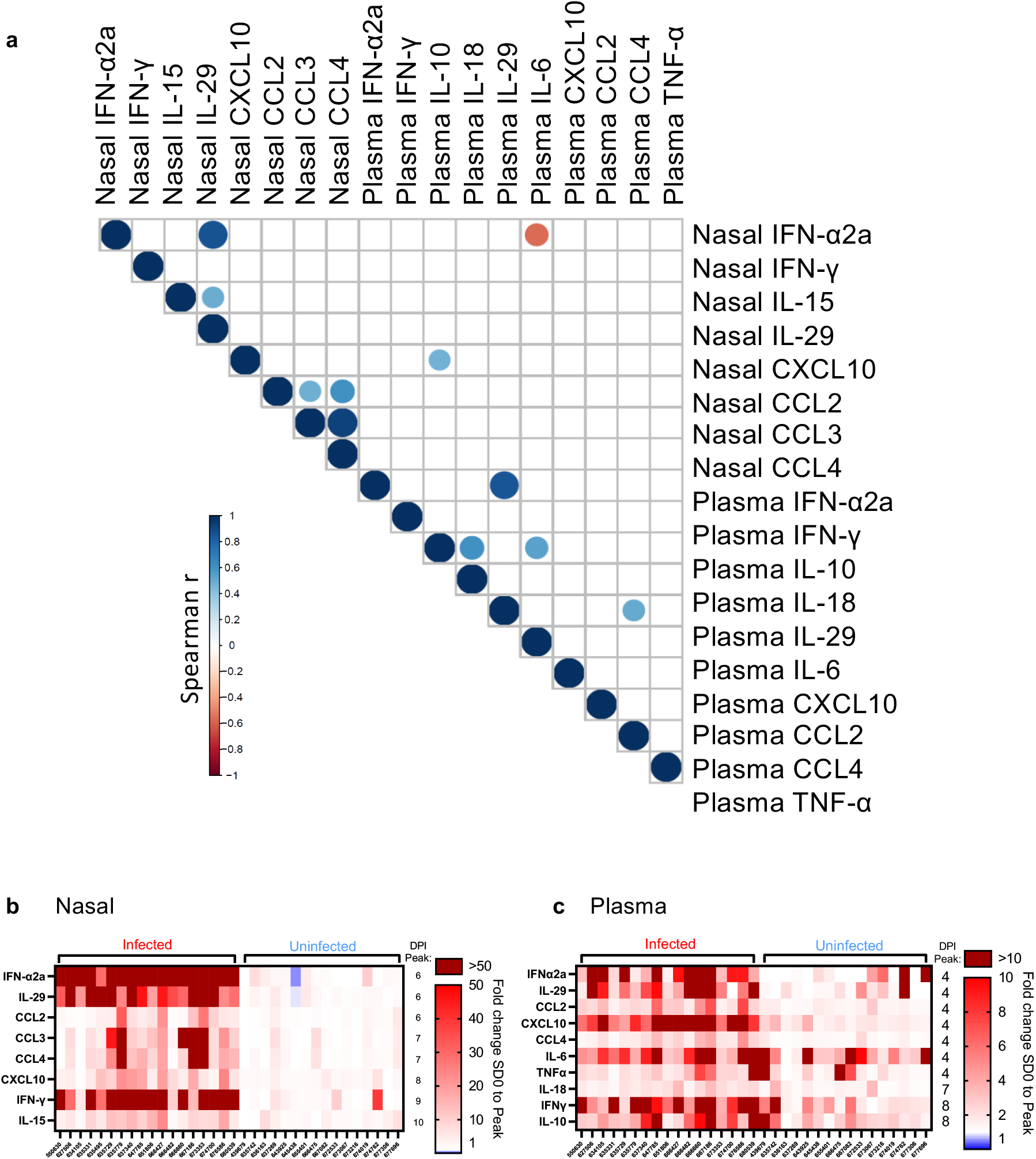
Correlations between local mucosal and systemic soluble mediator responses following SARS-CoV-2 inoculation. Correlation matrix shows significant (p = <0.05) Spearman r correlation between nasal and systemic (plasma) mediator responses (A). Heatmap of fold changes between baseline (day 0) and peak levels at any day post-inoculation in the infected and uninfected groups for nasal (B) or plasma (C) mediators shown.

**Supplementary Figure 3:**
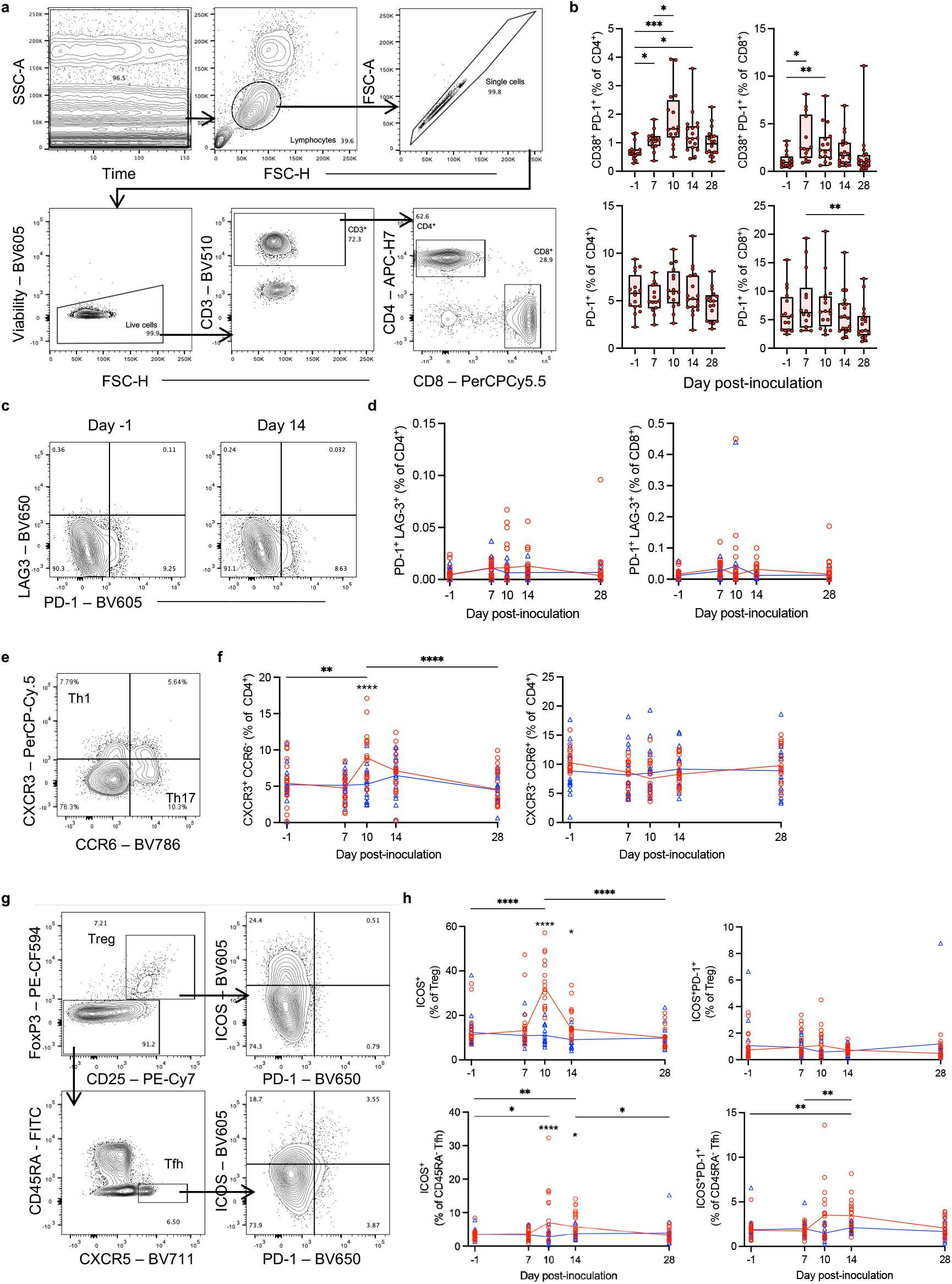
Extended phenotyping of the T cell response following SARS-CoV-2 inoculation. (A) Gating strategy for single, live, CD3^+^ lymphocytes and CD4^+^ or CD8^+^ T cells. (B) CD38^+^PD-1+ or PD-1 single positive T cells in the infected group (n=18) before and after SARS-CoV-2 inoculation. (C) Gating strategy for PD-1 and LAG-3 positive T cells and (D) frequencies of PD-1^+^LAG-3^+^ T cells in the infected (n=18) and uninfected (n=16) groups after inoculation. (E) Gating strategy for CXCR3 and CCR6 positive T cells and (F) frequencies of CXCR3^+^CCR6^-^ or CXCR3^-^CCR6^+^ T cells in the infected and uninfected groups after inoculation. (G) Gating strategy for FoxP3^+^CD25^+^ Treg and CD45RA^-^CXCR5^+^ cTfh T cells and ICOS and PD-1 expression Treg and cTfh T cells. (H) Frequencies of ICOS^+^ or ICOS^+^PD-1^+^ Treg or cTfh T cells in the infected and uninfected groups after inoculation. Participants vaccinated and infected in the community are excluded from day 28 data (see Supplementary Table 1 for details). Two-way ANOVA mixed-effects models with Geisser-Greenhouse correction for multiple testing was used to show significance between timepoints and groups. (*P* values *<0.05, **<0.01, ***<0.001, ****<0.0001).

**Supplementary Figure 4:**
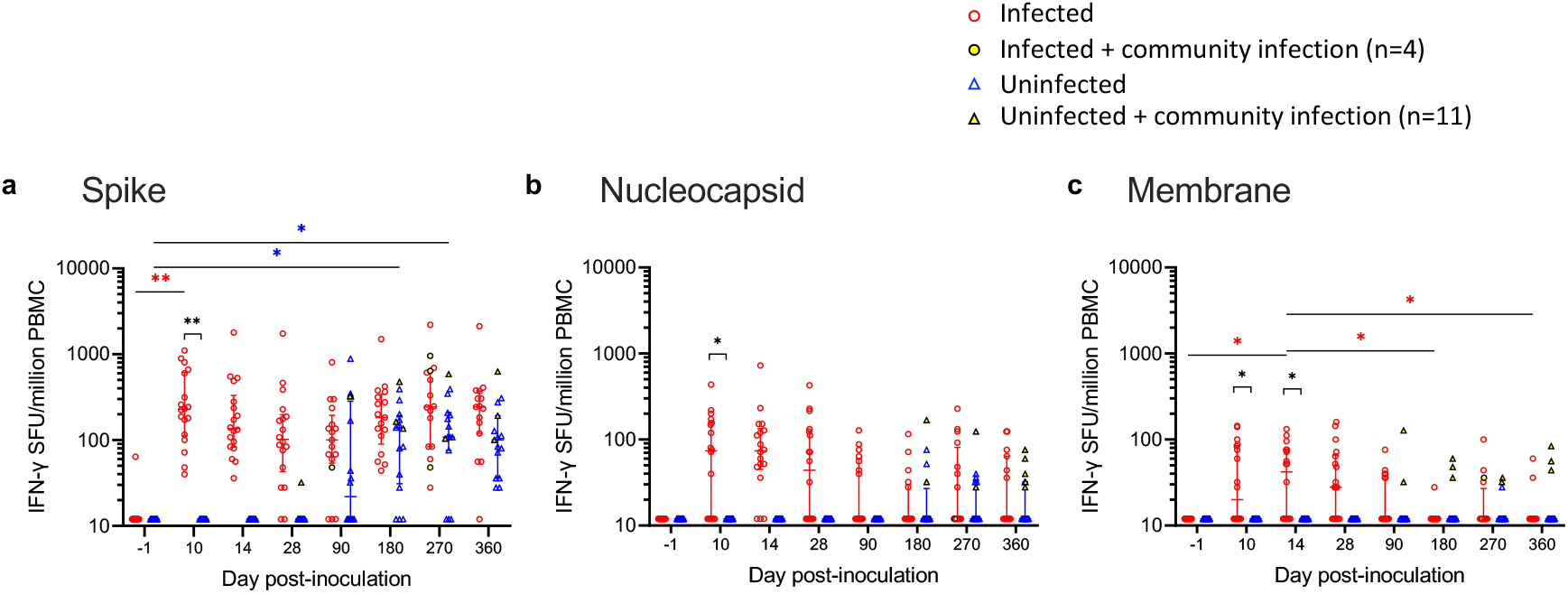
ELISpot responses following post-quarantine vaccination and infection in the community. ELISpot IFN-y spot forming units (SFU) per million PBMC are shown in response to (A) spike, (B) nucleocapsid and (C) membrane protein before and after inoculation up to day 360 in the infected group (n=18) and uninfected (n=16) group. All participants vaccinated and infected in the community are included at all timepoints. Two-way ANOVA mixed-effects models with Geisser-Greenhouse correction for multiple testing was used to show significance between timepoints and groups. (*P* values *<0.05, **<0.01, ***<0.001, ****<0.0001).

**Supplementary Figure 5:**
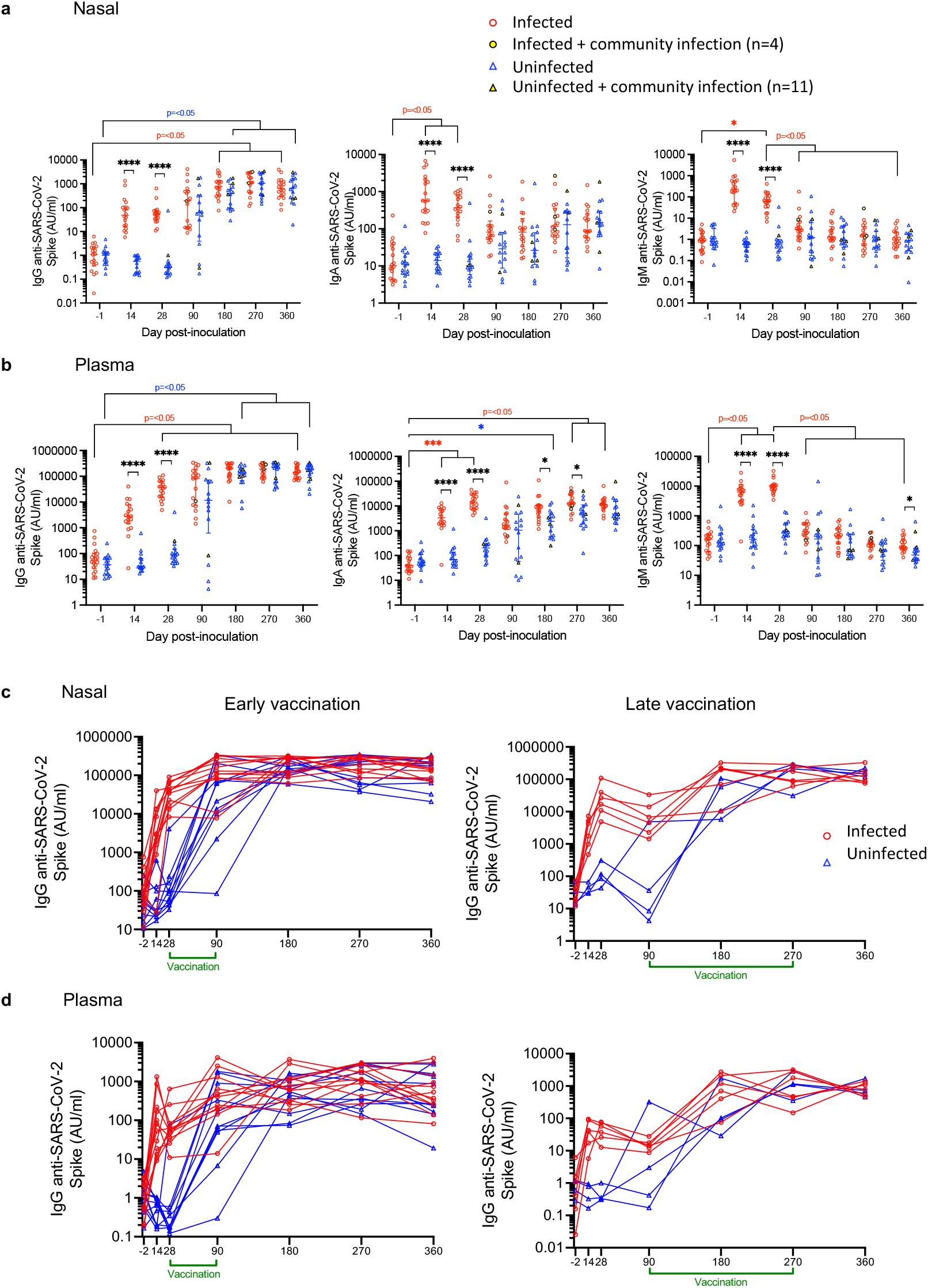
Antibody responses following post-quarantine vaccination and infection in the community. (A) Nasal and (B) plasma IgG, IgA and IgM anti-spike antibody responses measured by MSD before and after inoculation up to day 360 in the infected group (n=18) and uninfected (n=16) group are shown. All participants vaccinated and infected in the community are included at all timepoints. Anti-spike IgG in (C) the nose and (D) plasma is shown subdivided depending on whether the participant received the first SARS-CoV-2 vaccination dose early (day 28-90) or late (day 90-270). Two-way ANOVA mixed-effects models with Geisser-Greenhouse correction for multiple testing was used to show significance between timepoints and groups. (*P* values *<0.05, ***<0.001, ****<0.0001).

**Supplementary Figure 6:**
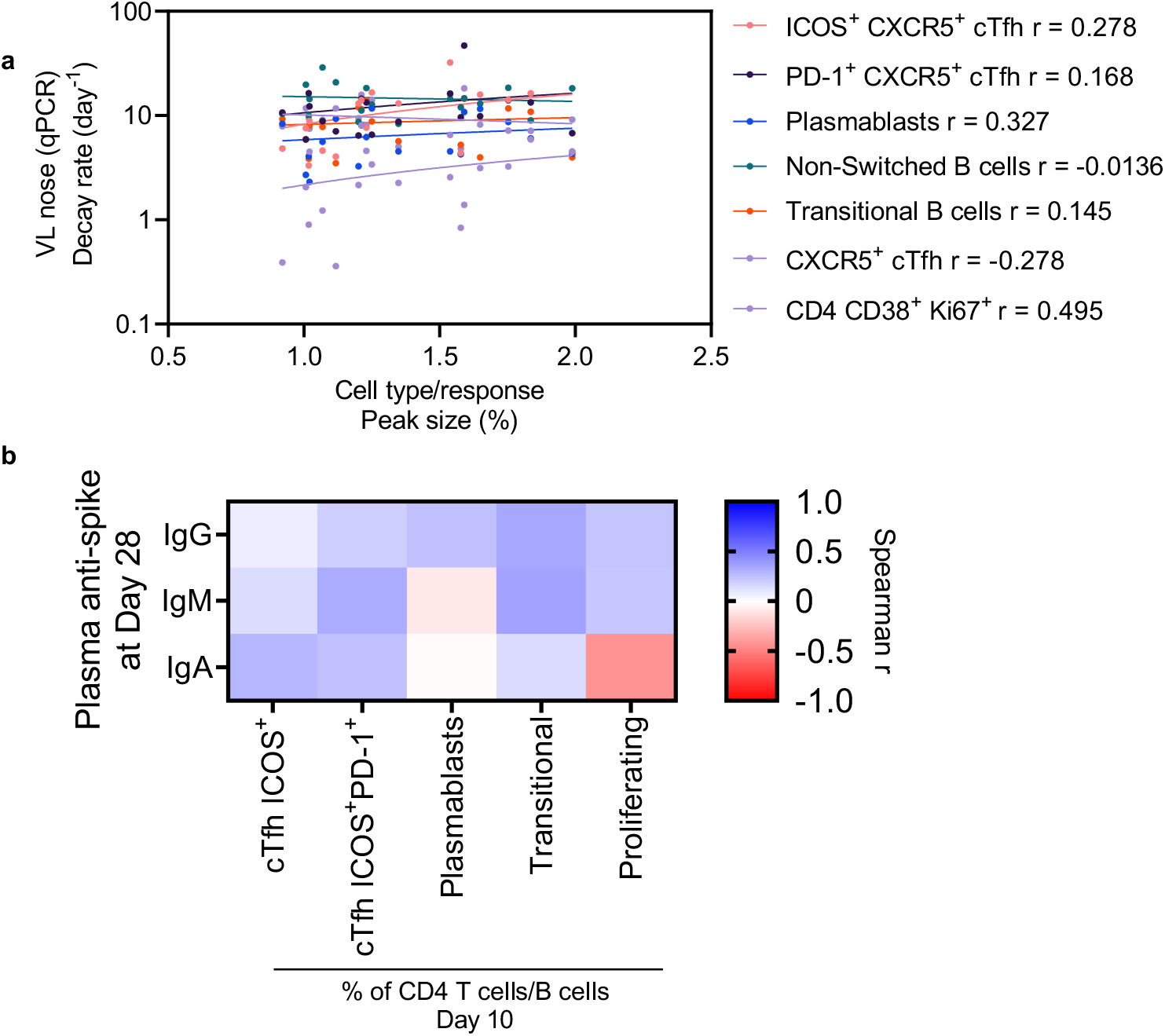
Extended data showing the lack of relationship between CD4^+^ T cell and B cell responses and viral load decline and antibody at day 28. (A) Correlation between activated and proliferating CD4^+^ T cells, cTfh T cells (including ICOS^+^ and PD-1^+^) or B cell subsets and VL decay rate. (B) Heat map showing the correlation between cTfh cell or B cell subsets and plasma antibody concentration at day 28 are shown (All ns).

**Supplementary Video 1:**
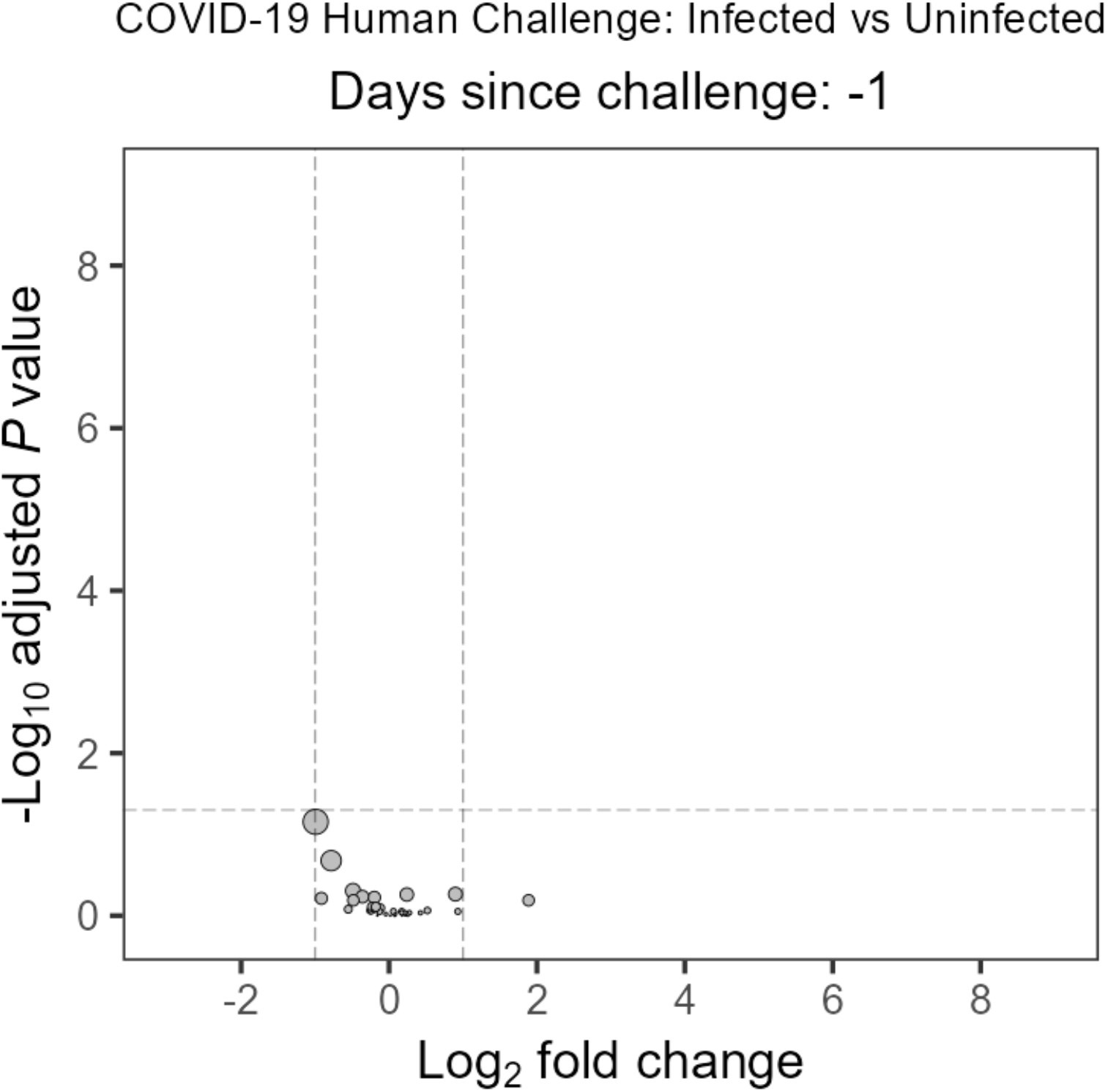
Soluble mediators in the nose were significantly different between infected and uninfected groups at different times over the study time course. Animated volcano plot showing Log_2_ fold change of geometric mean between infected and uninfected groups of soluble mediators measured in nasal lining fluid. Mann Whitney with Benjamini & Hochberg multiple comparison adjustment. Size of point = relative to effect size.

**Supplementary Table 1:** Vaccination and community infections in the post-quarantine period of the study (up to 1-year post-inoculation) and number of data points after exclusion at day 28 and 90.

|  | 1st vaccinations (total) | 1st vaccine dose day 14-28 | 1st vaccine dose day 28-90 | 1st vaccine dose day 90-180 | 1st vaccine dose day 180-270 |  |
| --- | --- | --- | --- | --- | --- | --- |
| Infected | 18 | 0 | 12 | 6 | 0 |  |
| Uninfected | 16<sup>a</sup> | 0 | 11 | 2 | 2 |  |
|  | Community infections (total) | 1st community infection day 14-28 | 1st community infection day 28-90 | 1st community infection day 90-180 | 1st community infection day 180-270 | 1st community infection day 270-360 |
| Infected | 4 | 0 | 1 | 0 | 3 | 0 |
| Uninfected | 13<sup>b</sup> | 1 | 5 | 2 | 3 | 2 |
|  | After excluding vaccinated/community infected, n= |  |  |  |  |  |
|  | Day 28 | Day 90 |  |  |  |  |
| Infected | 18 | 6 |  |  |  |  |
| Uninfected | 15 | 4 |  |  |  |  |

<sup>a</sup> One vaccination date unknown
<sup>b</sup> Two community infections were re-infections

