## Supplementary figures and images for "Mucosal and Systemic Immune Correlates of Viral Control following SARS-CoV-2 Infection Challenge in Seronegative Adults"

### Supplemental video 1

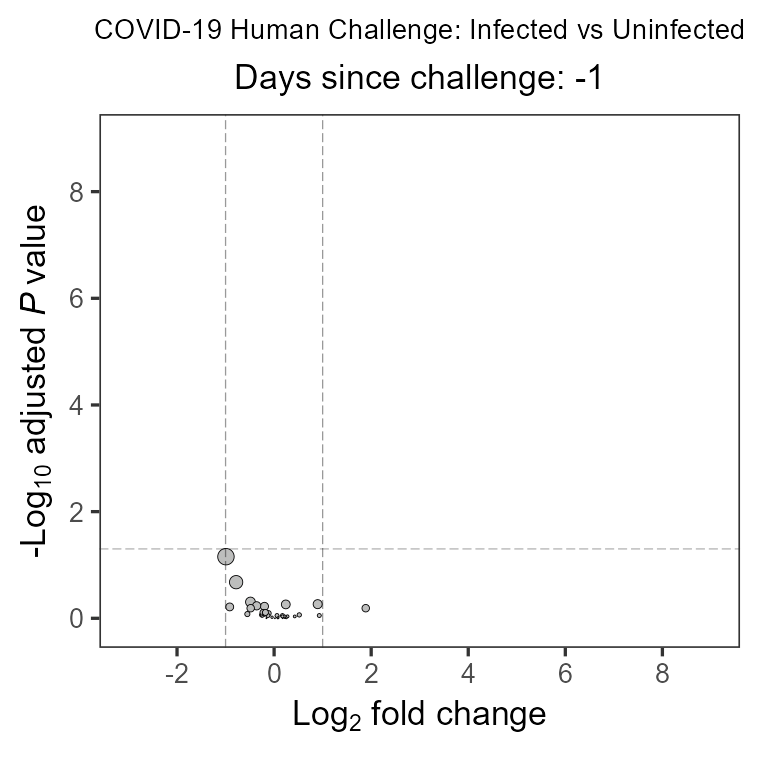
